# Robustness Analysis of Colorectal Cancer Colonoscopy Screening Strategies

**DOI:** 10.1101/2023.03.07.23286939

**Authors:** Pedro Nascimento de Lima, Carolyn M. Rutter, Christopher Maerzluft, Jonathan Ozik, Nicholson Collier

## Abstract

Colorectal Cancer (CRC) is a leading cause of cancer deaths in the United States. Despite significant overall declines in CRC incidence and mortality, there has been an alarming increase in CRC among people younger than 50. This study uses an established microsimulation model, CRC-SPIN, to perform a ‘stress test’ of colonoscopy screening strategies. First, we expand CRC-SPIN to include birth-cohort effects. Second, we estimate natural history model parameters via Incremental Mixture Approximate Bayesian Computation (IMABC) for two model versions to characterize uncertainty while accounting for increased early CRC onset. Third, we simulate 26 colonoscopy screening strategies across the posterior distribution of estimated model parameters, assuming four different colonoscopy sensitivities (104 total scenarios). We find that model projections of screening benefit are highly dependent on natural history and test sensitivity assumptions, but in this stress test, the policy recommendations are robust to the uncertainties considered.

## 1 Background

Colorectal Cancer (CRC) is the second-leading cause of cancer deaths in the United States. In 2021, 104,270 new CRC cases and 52,980 CRC cancer deaths were projected to occur in the US (Siegel et al. 2021). From 1991 through 2018, cancer death rates fell by 31%, a consistent decline attributed to smoking reduction, early detection, and improved treatment. For CRC specifically, the decrease in annual mortality from 1980 through 2018 was 53% among males (from 32.8 to 15.8 annual deaths per 100,000 people) and 55% among females (from 24.4 to 10.9 annual deaths per 100,000 people) (NCI 2021).

Despite significant overall decreases in CRC incidence, CRC incidence has increased among adults younger than 50. Adults born around 1990 have double the risk of colon cancer and quadruple the risk of rectal cancer compared to those born in 1950 (Siegel et al. 2017). This concerning development has raised questions about the potential causes of this phenomenon and the appropriate policy responses. While an Age-Period-Cohort model (Siegel et al. 2017) identified a birth cohort effect underlying the increase in incidence, there is no consensus on the specific causes. Current screening guidelines have reduced the recommended age to start screening from 50 to 45 years (Knudsen et al. 2021a).

To date, CRC microsimulation models do not incorporate birth cohort effects. The absence of an integrated approach to account for changing risk may be problematic for several reasons. First, the data used to calibrate microsimulation models spans several decades. If the natural history of the disease has changed over time, those changes cause confounding in parameter estimates and tensions between calibration targets that might be reconciled if models were extended to include birth cohort effects. Second, data from new studies may conflict with existing model predictions that do not address changes in risk. The absence of birth cohort effects may undermine the validation of existing models if those effects prove consequential. Finally, the hypothesis of no birth cohort effects is inconsistent with recent cancer incidence data (Siegel et al. 2017).

This analysis informs the debate around policy responses to the early onset of CRC by evaluating the robustness of currently recommended (Knudsen et al. 2021a) CRC screening policies to a set of uncertainties. First, we perform Bayesian calibration of two specifications of the CRC-SPIN model (Rutter and Savarino 2010) to characterize both structural and parametric uncertainties surrounding the natural history of CRC. Both model specifications include a non-parametric birth cohort effects model. Second, we simulate all colonoscopy screening strategies evaluated in the cost-effectiveness analysis that informed the most recent USTFPS screening recommendations (Knudsen et al. 2021a) to assess how screening cost-effectiveness depends on within and between those models. Finally, we investigate whether current colonoscopy screening recommendations are robust to uncertainties surrounding the natural history of CRC and the sensitivity of colonoscopy by investigating the stability of the cost-effectiveness frontier.

## 2 Methods

### 2.1 Natural History Model

#### 2.1.1 Adenoma Risk with Birth Cohort Effects

CRC-SPIN simulates adenoma risk using a Non-homogenous Poisson Process, as shown in eq. (1). Individual log-risk Ψ_ia_ of person i at age a depends on their sex, age, and birth year. This equation includes two modifications relative to previous CRC-SPIN versions. First, eq. (1) allows the specification of adenoma risk change points *k*_0_, *k*_1_, *k*_2_, *k*_3_ instead of fixed change points previously used in the baseline model (*k*_0_ = 20, *k*_1_ = 50, *k*_2_ = 60, *k*_3_ = 70). Second, the model allows for increased adenoma initiation risk based on the birth year of each individual. In eq. (1), δ(*x*) = 1 when *x* is true and 0 otherwise. *α*_0i_ ∼ *N*(*A*_a_, *σ*_a_) allows for between-individual variation in adenoma risk. All risk parameters *A*_a_, *σ*_a_, *α*_1_, *α*_2_, *α*_*k_0_*_, *α*_*k_1_*_, *α*_*k_2_*_, and *α*_*k_3_*_ are unknown parameters that need estimation.

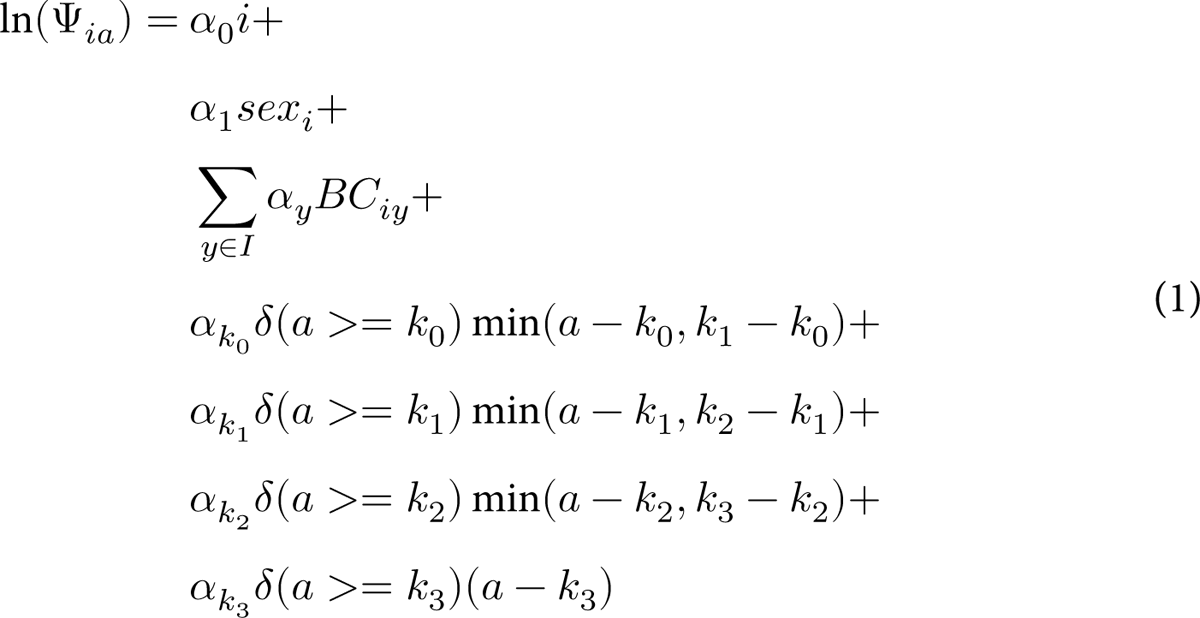

The birth cohort effects model defines adenoma risk birth-cohort effects *α*_y_ for each birth-cohort year within the set of years *I* = [1880, 1881, …, 1975], allowing the population-level average risk to change. This range of years encompasses the range of birth cohorts represented by calibration targets used by the model and can be expanded or contracted as information from new birth cohorts become available. The indicator variable *BC*_iy_ is one if individual *i* was born in year *y* and zero otherwise. This effect captures secular changes that resulted in higher adenoma risk while making no additional assumptions about the underlying causes and minimal assumptions about the functional form of those changes.

Because a non-parametric model of birth cohort effects with free year parameters substantially expands the parameter space, we use the following strategy to enable the parameter search while allowing a flexible functional form. First, we define a set of years in which the birth cohort effect will be estimated: 1980, 1910, 1940, 1955, 1970, and 1975. The year 1940 is a reference point at which the effect is zero. The minimum and maximum dates are chosen based on the available calibration targets. The intermediate knots were chosen to be evenly spaced, with a finer resolution after 1940, where risk changes were known to be pronounced. Second, we produce a smooth, monotonic, biologically-plausible interpolation between these points to define birth cohort effects between the years used as knots, using a method based on piecewise radial functions (Stineman 1980; Jhannesson, Bjornsson, and Grothendieck 2018)^1^. Finally, we enforce monotonic increases in risk after 1950, following the evidence found in Age-Period-Cohort analyses (Siegel et al. 2017). After initiation, adenomas are distributed in the large intestine following a multinomial distribution with probabilities *P (l = Rectum) = 0.09, P (l = Sigmoid Colon) = 0.24, P (Descending Colon) = 0.12, P (l = TransverseColon) = 0.24, P (l = Ascending Colon) = 0.23, and P (l = Cecum) = 0.08*.

#### 2.1.2 Model Specifications

This paper employs two alternative specifications of the CRC-SPIN model. In the *baseline model specification (BC-20)*, we set the minimum age at adenoma initiation *k*_0_ R 20, consistent with prior CRC-SPIN analyses. In a second model specification *(BC-10)*, we set *k*_0_ R 10, allowing adenomas to be initiated after age 10. The age at which adenomas are initiated can be considered a *deep uncertainty* because there is little information to estimate this parameter. Increasing cancer incidence at younger ages (Siegel et al. 2017) might suggest that a model with a younger minimum age at first adenoma initiation, at age 10, could be more plausible than a model with an older minimum age of 20. We investigate this question by calibrating and evaluating screening policies’ robustness across both model specifications. This approach can be readily extended to more than two model specifications. The only reason we do not present additional model specifications is the computational burden of adding a new model to this analysis.

### 2.2 Bayesian Inference of Natural History Parameters

Approximate Bayesian Computation (ABC) is an inference framework that requires the specification of i) calibration targets defined as summary statistics derived from data, ii) prior distributions for a set of unknown parameters, and iii) a model that maps parameter sets to model-predicted targets. We use the Incremental Mixture Approximate Bayesian Computation (IMABC) algorithm (Rutter, Ozik, DeYoreo, et al. 2019) to estimate the natural history model’s joint distribution of parameters. Based on these three elements, IMABC iteratively samples the parameter space and approximates the posterior distribution of model parameters. Table 1 presents the prior distribution for each parameter used across both model specifications.

**Table 1:**
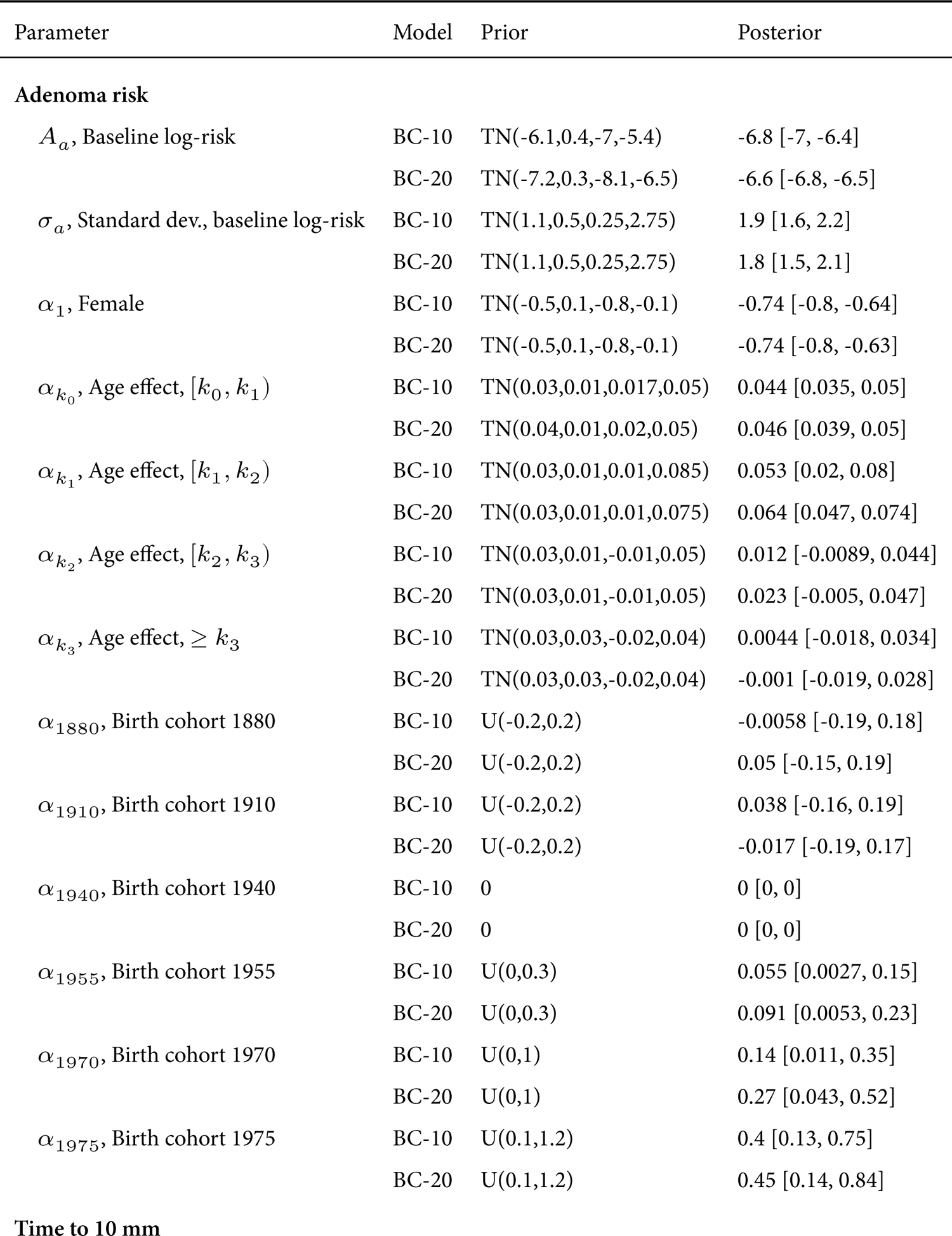
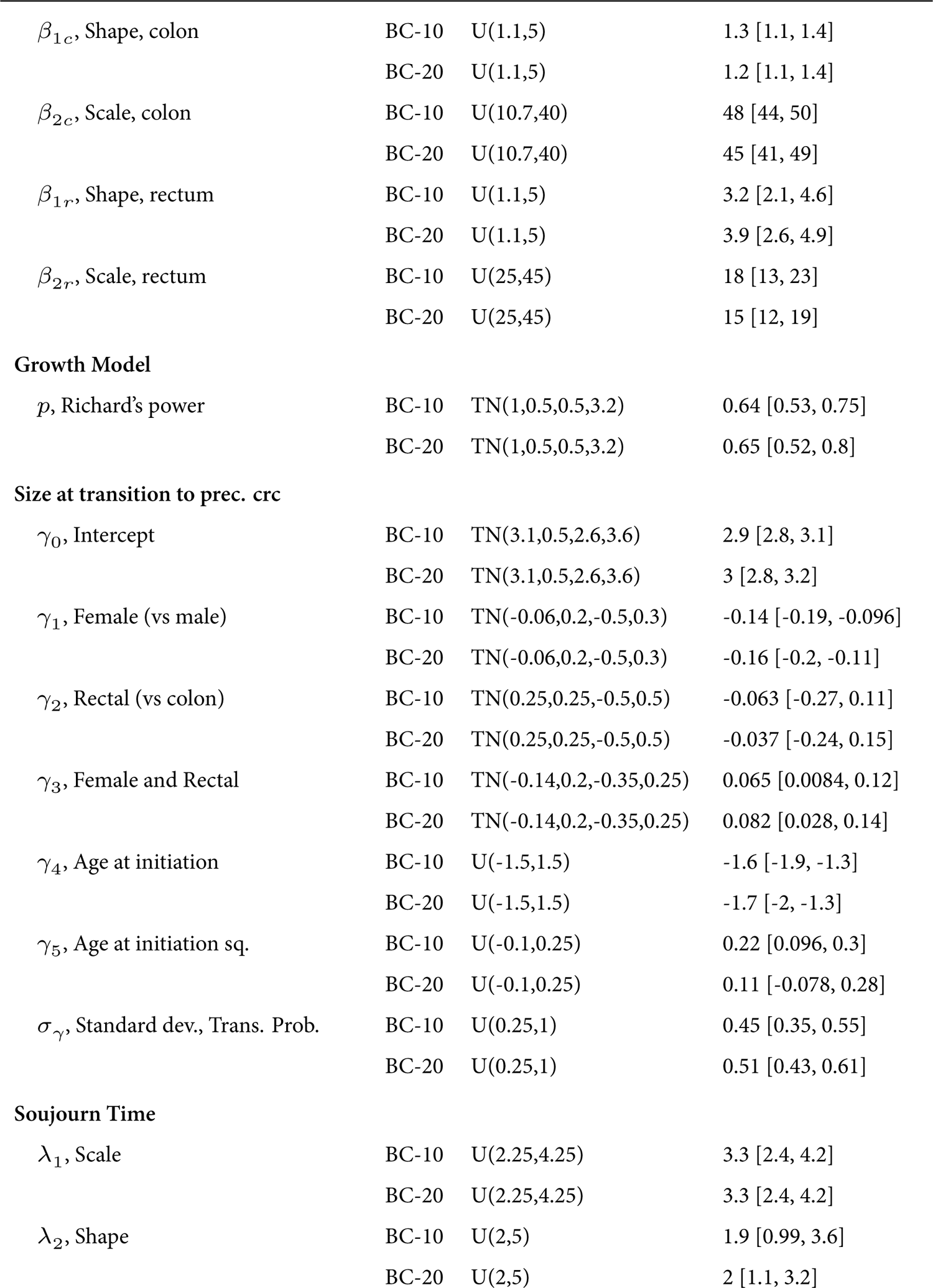
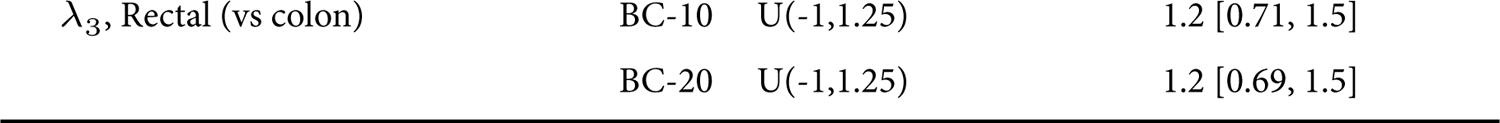
Parameter Prior and Posterior Distributions

| Parameter | Model | Prior | Posterior |
| --- | --- | --- | --- |
| **Adenoma risk** |  |  |  |
| $A_a$, Baseline log-risk | BC-10 | TN(-6.1,0.4,-7,-5.4) | -6.8 [-7, -6.4] |
|  | BC-20 | TN(-7.2,0.3,-8.1,-6.5) | -6.6 [-6.8, -6.5] |
| $\sigma_a$, Standard dev., baseline log-risk | BC-10 | TN(1.1,0.5,0.25,2.75) | 1.9 [1.6, 2.2] |
|  | BC-20 | TN(1.1,0.5,0.25,2.75) | 1.8 [1.5, 2.1] |
| $\alpha_1$, Female | BC-10 | TN(-0.5,0.1,-0.8,-0.1) | -0.74 [-0.8, -0.64] |
|  | BC-20 | TN(-0.5,0.1,-0.8,-0.1) | -0.74 [-0.8, -0.63] |
| $\alpha_{k_0}$, Age effect, $[k_0, k_1)$ | BC-10 | TN(0.03,0.01,0.017,0.05) | 0.044 [0.035, 0.05] |
|  | BC-20 | TN(0.04,0.01,0.02,0.05) | 0.046 [0.039, 0.05] |
| $\alpha_{k_1}$, Age effect, $[k_1, k_2)$ | BC-10 | TN(0.03,0.01,0.01,0.085) | 0.053 [0.02, 0.08] |
|  | BC-20 | TN(0.03,0.01,0.01,0.075) | 0.064 [0.047, 0.074] |
| $\alpha_{k_2}$, Age effect, $[k_2, k_3)$ | BC-10 | TN(0.03,0.01,-0.01,0.05) | 0.012 [-0.0089, 0.044] |
|  | BC-20 | TN(0.03,0.01,-0.01,0.05) | 0.023 [-0.005, 0.047] |
| $\alpha_{k_3}$, Age effect, $\geq k_3$ | BC-10 | TN(0.03,0.03,-0.02,0.04) | 0.0044 [-0.018, 0.034] |
|  | BC-20 | TN(0.03,0.03,-0.02,0.04) | -0.001 [-0.019, 0.028] |
| $\alpha_{1880}$, Birth cohort 1880 | BC-10 | U(-0.2,0.2) | -0.0058 [-0.19, 0.18] |
|  | BC-20 | U(-0.2,0.2) | 0.05 [-0.15, 0.19] |
| $\alpha_{1910}$, Birth cohort 1910 | BC-10 | U(-0.2,0.2) | 0.038 [-0.16, 0.19] |
|  | BC-20 | U(-0.2,0.2) | -0.017 [-0.19, 0.17] |
| $\alpha_{1940}$, Birth cohort 1940 | BC-10 | 0 | 0 [0, 0] |
|  | BC-20 | 0 | 0 [0, 0] |
| $\alpha_{1955}$, Birth cohort 1955 | BC-10 | U(0,0.3) | 0.055 [0.0027, 0.15] |
|  | BC-20 | U(0,0.3) | 0.091 [0.0053, 0.23] |
| $\alpha_{1970}$, Birth cohort 1970 | BC-10 | U(0,1) | 0.14 [0.011, 0.35] |
|  | BC-20 | U(0,1) | 0.27 [0.043, 0.52] |
| $\alpha_{1975}$, Birth cohort 1975 | BC-10 | U(0.1,1.2) | 0.4 [0.13, 0.75] |
|  | BC-20 | U(0.1,1.2) | 0.45 [0.14, 0.84] |
| **Time to 10 mm** |  |  |  |

Table 1: Parameter Prior and Posterior Distributions (*continued*)
| Parameter | Model | Prior | Posterior |
| --- | --- | --- | --- |
| $\beta_{1c}$, Shape, colon | BC-10 | U(1.1,5) | 1.3 [1.1, 1.4] |
|  | BC-20 | U(1.1,5) | 1.2 [1.1, 1.4] |
| $\beta_{2c}$, Scale, colon | BC-10 | U(10.7,40) | 48 [44, 50] |
|  | BC-20 | U(10.7,40) | 45 [41, 49] |
| $\beta_{1r}$, Shape, rectum | BC-10 | U(1.1,5) | 3.2 [2.1, 4.6] |
|  | BC-20 | U(1.1,5) | 3.9 [2.6, 4.9] |
| $\beta_{2r}$, Scale, rectum | BC-10 | U(25,45) | 18 [13, 23] |
|  | BC-20 | U(25,45) | 15 [12, 19] |
| **Growth Model** |  |  |  |
| $p$, Richard's power | BC-10 | TN(1,0.5,0.5,3.2) | 0.64 [0.53, 0.75] |
|  | BC-20 | TN(1,0.5,0.5,3.2) | 0.65 [0.52, 0.8] |
| **Size at transition to prec. crc** |  |  |  |
| $\gamma_0$, Intercept | BC-10 | TN(3.1,0.5,2.6,3.6) | 2.9 [2.8, 3.1] |
|  | BC-20 | TN(3.1,0.5,2.6,3.6) | 3 [2.8, 3.2] |
| $\gamma_1$, Female (vs male) | BC-10 | TN(-0.06,0.2,-0.5,0.3) | -0.14 [-0.19, -0.096] |
|  | BC-20 | TN(-0.06,0.2,-0.5,0.3) | -0.16 [-0.2, -0.11] |
| $\gamma_2$, Rectal (vs colon) | BC-10 | TN(0.25,0.25,-0.5,0.5) | -0.063 [-0.27, 0.11] |
|  | BC-20 | TN(0.25,0.25,-0.5,0.5) | -0.037 [-0.24, 0.15] |
| $\gamma_3$, Female and Rectal | BC-10 | TN(-0.14,0.2,-0.35,0.25) | 0.065 [0.0084, 0.12] |
|  | BC-20 | TN(-0.14,0.2,-0.35,0.25) | 0.082 [0.028, 0.14] |
| $\gamma_4$, Age at initiation | BC-10 | U(-1.5,1.5) | -1.6 [-1.9, -1.3] |
|  | BC-20 | U(-1.5,1.5) | -1.7 [-2, -1.3] |
| $\gamma_5$, Age at initiation sq. | BC-10 | U(-0.1,0.25) | 0.22 [0.096, 0.3] |
|  | BC-20 | U(-0.1,0.25) | 0.11 [-0.078, 0.28] |
| $\sigma_\gamma$, Standard dev., Trans. Prob. | BC-10 | U(0.25,1) | 0.45 [0.35, 0.55] |
|  | BC-20 | U(0.25,1) | 0.51 [0.43, 0.61] |
| **Soujourn Time** |  |  |  |
| $\lambda_1$, Scale | BC-10 | U(2.25,4.25) | 3.3 [2.4, 4.2] |
|  | BC-20 | U(2.25,4.25) | 3.3 [2.4, 4.2] |
| $\lambda_2$, Shape | BC-10 | U(2,5) | 1.9 [0.99, 3.6] |
|  | BC-20 | U(2,5) | 2 [1.1, 3.2] |

Table 1: Parameter Prior and Posterior Distributions (*continued*)
| Parameter | Model | Prior | Posterior |
| --- | --- | --- | --- |
| $\lambda_3$, Rectal (vs colon) | BC-10 | U(-1,1.25) | 1.2 [0.71, 1.5] |
|  | BC-20 | U(-1,1.25) | 1.2 [0.69, 1.5] |

#### 2.2.1 Calibration Targets

This analysis uses a set of 43 calibration targets derived from eight sources to calibrate CRC-SPIN. Following prior CRC-SPIN calibrations (Rutter, Ozik, Deyoreo, et al. 2019; DeYoreo et al. 2022), we use data from Corley et al. (2013) to inform adenoma prevalence by sex and age. Pickhardt et al. (2003) informs the distribution of adenoma size. Church (2004) and Lieberman et al. (2008) inform the distribution of CRC size. The UK Flexible Sigmoidoscopy Screening Trial (Atkin et al. 2010) informs the proportion of individuals who are screen-detected with CRC by sex. Cancer incidence data by sex, age, and location (rectal vs. colon) from SEER 1975-1979 (National Cancer Institute 2022) informs cancer incidence risk.

This analysis included two new targets relative to prior CRC-SPIN calibrations. Increased risk among new cohorts is informed by recent SEER data (National Cancer Institute 2022), using SEER 2009 and SEER 2014 incidence for young adults (aged 40-44). Table 2 lists each calibration target, tolerance intervals used in this calibration, and the mean and 95% credible intervals of the posterior distribution obtained from calibration.

**Table 2:** Calibration Targets and Tolerance Intervals

| Name | Target (Tolerance Interval) |
| --- | --- |
| **Corley (2013)** |  |
| Adenoma prevalence, male [50 – 54] | 25 (21.2,35) |
| Adenoma prevalence, male [55 – 59] | 29 (24.6,40.6) |
| Adenoma prevalence, male [65 – 64] | 31 (26.4,43.4) |
| Adenoma prevalence, male [65 – 69] | 34 (28.8,48.3) |
| Adenoma prevalence, male [70 – 74] | 39 (32,56.8) |
| Adenoma prevalence, male $\geq 75$ | 38 (30.4,56.3) |
| Adenoma prevalence, female [50 – 54] | 15 (12.8,21.1) |
| Adenoma prevalence, female [55 – 59] | 18 (15.3,25.4) |
| Adenoma prevalence, female [65 – 64] | 22 (18.7,30.8) |
| Adenoma prevalence, female [65 – 69] | 24 (20.2,34.2) |
| Adenoma prevalence, female [70 – 74] | 26 (21.1,38.1) |
| Adenoma prevalence, female $\geq 75$ | 26 (20.4,39.1) |
| **UKFSS, Atkin (2010)** |  |
| Detected preclinical CRC per 1000 people , male | 4.68 (3.26,6.47) |
| Detected preclinical CRC per 1000 people ,female | 1.74 (0.93,2.93) |
| **SEER 1975-1979, NCI (2012)** |  |
| CRC incidence, colon, female [40 – 49] | 14.8 (11.8,17.8) |
| CRC incidence, colon, female [50 – 59] | 43.2 (36.8,49.7) |
| CRC incidence, colon, female [60 – 69] | 101 (86,116) |
| CRC incidence, colon, female [70 – 84] | 219 (186,252) |
| CRC incidence, colon, female $\geq 85$ | 295 (250,339) |
| CRC incidence, colon, male [40 – 49] | 13.7 (10.7,16.7) |
| CRC incidence, colon, male [50 – 59] | 45.8 (38.9,52.7) |
| CRC incidence, colon, male [60 – 69] | 123 (104,141) |
| CRC incidence, colon, male [70 – 84] | 273 (232,314) |
| CRC incidence, colon, male $\geq 85$ | 386 (328,444) |
| CRC incidence, rectal, female [40 – 49] | 5.84 (2.84,8.84) |
| CRC incidence, rectal, female [50 – 59] | 20.5 (17.4,23.6) |
| CRC incidence, rectal, female [60 – 69] | 43.2 (36.7,49.6) |

Table 2: Calibration Targets and Tolerance Intervals (*continued*)
| Name | Target (Tolerance Interval) |
| --- | --- |
| CRC incidence, rectal, female [70 – 84] | 75.6 (64.3,87) |
| CRC incidence, rectal, female $\geq 85$ | 104 (88.2,119) |
| CRC incidence, rectal, male [40 – 49] | 7.23 (4.23,10.2) |
| CRC incidence, rectal, male [50 – 59] | 30.2 (25.6,34.7) |
| CRC incidence, rectal, male [60 – 69] | 72.4 (61.6,83.3) |
| CRC incidence, rectal, male [70 – 84] | 131 (111,150) |
| CRC incidence, rectal, male $\geq 85$ | 164 (139,190) |
| **SEER 2009-2014, NCI (2012)** |  |
| CRC incidence, [40 – 44] | 16.4 (13.4,19.4) |
| CRC incidence, [40 – 44] | 14.4 (11.4,17.4) |
| **Pickhardt (2003)** |  |
| Percent of detected adenomas $< 6$ mm | 62.1 (52.8,71.4) |
| Percent of detected adenomas [6, 10) mm | 28.7 (22.6,35.4) |
| Percent of detected adenomas $\geq 10$mm | 9.21 (5.64,13.9) |
| **Church (2004)** |  |
| Preclinical CRCs per 1000 lesions [6, 10) mm | 2.39 (0.0012,23.7) |
| Preclinical CRCs per 1000 lesions $\geq 10$mm | 42.3 (18.5,80.6) |
| **Lieberman (2008)** |  |
| Preclinical CRCs per 1000 lesions [6, 10) mm | 2.51 (0.0402,15) |
| Preclinical CRCs per 1000 lesions $\geq 10$mm | 32.8 (15.5,59.7) |

#### 2.2.2 Incorporating New Information via Sequential Calibration

This study used the following strategy to calibrate birth cohort effect parameters. For each candidate parameter set evaluated by the model, we first evaluate targets that are fast to simulate (e.g., Corley et al. (2013) adenoma prevalence) and only evaluate computationally-expensive targets (SEER cancer incidence) for parameters that result in adequate predictions of these easy-to-simulate targets. This approach leverages IMABC’s iterative tolerance interval updating strategy and saves computing time by not spending resources on parameter sets that could not have generated the calibration target data.

After calibrating the model to targets used in previous analyses (Rutter, Ozik, DeYoreo, et al. 2019), we incorporated recent information from CRC incidence using a sequential approach. As demonstrated by DeYoreo et al. (2022), one can add new information to the model by adding calibration targets after calibrating the model to an initial set of targets. First, we used IMABC to calibrate 41 calibration targets, except for the two SEER cancer incidence targets that contain new information about increased CRC risk in recent cohorts (SEER 2009 and SEER 2014). Because these two targets use approximately 1/3 of the computing time required to simulate all targets, avoiding evaluating them before all other targets are met allowed the algorithm to build a posterior distribution without the burden of new, expensive targets. After building a posterior distribution with an effective sample size (ESS) greater than 10,000 for each model specification, we evaluated the new targets across this distribution and selected the parameters within all tolerance intervals for all calibration targets. This second step is akin to a one-step rejection sampling ABC scheme, wherein we use the posterior of prior calibration to condition the parameter distribution on new data. This approach is very similar to the sequential approach used by DeYoreo et al. (2022), with the exception that we do not need to perform new IMABC iterations to increase the size of the new parameter distribution because the resulting distribution is already dense enough (sample size > 1000) to support the analysis of screening strategies.

### 2.3 Colonoscopy Screening Strategies

We used the two calibrated birth cohort models to project outcomes for the 26 colonoscopy screening strategies evaluated in existing CRC screening recommendations (Knudsen et al. 2021a). These strategies involve three policy levers: age to start screening (45, 50, 55), periodicity of screening colonoscopies (5, 10, 15), and age to end screening (70, 75, 80, 85). A grid experimental design over the three policy levers results in 36 strategies, but only 26 are simulated after removing combinations resulting in redundant strategies. Screening strategies are coded as *[age to start]-[age to end],[interval]*. For instance, strategy *45-75,10* refers to performing colonoscopy screening at ages 45, 55, 65, and 75. Following comparable studies (Knudsen et al. 2021a), perfect adherence to screening is assumed across all model runs.

#### 2.3.1 Colonoscopy Sensitivity Scenarios

The sensitivity of colonoscopy exams is uncertain and very heterogeneous across gastroenterologists and gastroenterology clinics (Corley et al. 2014). Hence, assessing how sensitivity affects the expected benefit from different screening strategies is important. Table 3 presents the sensitivity assumptions made in four sensitivity scenarios considered in this analysis and corresponding sources. Section 8 provides an extended discussion around the rationale for considering those scenarios and evaluates the plausibility of the “Very Low” sensitivity scenario by re-analyzing data from a meta-analysis of tandem colonoscopy studies (Van Rijn et al. 2006). This analysis assumes time-independent, uncorrelated colonoscopy sensitivity conditional on lesion size and no heterogeneity in sensitivity across the population.

**Table 3:** Colonoscopy Sensitivity Assumtpions

| Adenoma Size | Sensitivity | Source |
| --- | --- | --- |
| **Very Low** |  |  |
| <=5 mm | 0.550 | Appendix II |
| 6-9 mm | 0.700 | Rutter (2021) |
| >= 10 mm | 0.900 | Rutter (2021) |
| **Low** |  |  |
| <=5 mm | 0.700 | Zauber (2008) |
| 6-9 mm | 0.800 | Zauber (2008) |
| >= 10 mm | 0.931 | Knudsen (2016) |
| **Baseline** |  |  |
| <=5 mm | 0.750 | Zauber (2008) |
| 6-9 mm | 0.850 | Zauber (2008) |
| >= 10 mm | 0.950 | Zauber (2008) |
| **High** |  |  |
| <=5 mm | 0.790 | Zauber (2008) |
| 6-9 mm | 0.920 | Zauber (2008) |
| >= 10 mm | 0.995 | Knudsen (2016) |

#### 2.3.2 Cost-Effectiveness Outcomes

This study simulates a cohort of 2 million average-risk 40-year-old adults from the general US population followed through death. Following recent analyses used to guide policy (Knudsen et al. 2021a), we use Life-years Gained (*LYG*) from screening as the effectiveness measure by comparing a no-screening scenario to each scenario where individuals are screened. We perform this comparison for each parameter set and population block. The number of colonoscopies (*n*_col_) performed is used as the burden measure. Incremental efficiency ratios (ICER)^2^ are computed by comparing two non-dominated screening strate-gies as 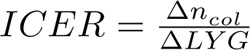 where Δ*n_col_* is the additional number of colonoscopies required by a more intensive screening regimen and *ΔLYG* is the incremental number of life-years gained by that regimen. We also remove extended-dominated strategies from the Pareto frontier so that efficiency ratios increase monotonically with effectiveness.

#### 2.3.3 Experimental Design

Following the Robust Decision Making (RDM) decision-analytic approach (Lempert et al. 2006), we created a large-scale experimental design to stress-test the cost-effectiveness of alternative screening strategies. The experimental design of this study considered natural history uncertainty (represented by the posterior distribution of model parameters), structural assumptions (represented by two different model specifications), and uncertainty related to the efficacy of the interventions (represented by the four sensitivity scenarios). Hence, the experimental design of this study consisted of the combination of 2 model specifications, 500 natural history parameter sets for each model specification, 26 screening strategies and 1 “No-Screening” scenario used as the comparator, and 4 colonoscopy sensitivity scenarios, resulting in 105,000 unique model runs. The dimensions of the experimental design were defined such that the screening experiments would be performed using a computing budget of approximately 200,000 core hours. Each unique model run simulates 2 million individuals^3^. This experimental design results in 840 model runs in this experiment, which results in 210 billion life histories and over 0.63 trillion adenomas.

### 2.4 High-performance Computing Infrastructure

The scale of this experiment required the design and development of high-performance computing (HPC) tools and workflows. Experiments were run on computing resources at Argonne’s Laboratory Computing Resource Center and the Argonne Leadership Computing Facility. The model code for this experiment was developed in R, and HPC workflows were developed using Swift-T (Wozniak et al. 2013), EMEWS (Ozik et al. 2016), and the crcrdm R package (Nascimento de Lima 2022). These experiments also used a relational Postgres SQL database to allow for massively parallel execution of experiments and rapid retrieval of results. More information about the computational environment and software developed for these experiments can be found in sections 9 and 10.

## 3 Results

### 3.1 Natural History Parameter Estimates

Appendix table 1 presents model parameters, their prior distribution used for calibration, and their respective 95% posterior credible intervals for each model. While some parameter estimates are similar across models, these results demonstrate that model estimates are contingent on the assumed minimum age at adenoma initiation. For instance, model All estimates were presented with up to three significant digits of precision. BC-20 predicts lower adenoma prevalence at age 25 than model BC-10, and the posterior distributions of the models do not overlap. Moreover, the minimum age at which adenomas are allowed to start also affects the estimates of birth cohort effects. If adenomas are initiated earlier (as in model BC-10), then the birth cohort effects needed to explain increased incidence are also expected to be lower.

While table 1 presents a high-level summary of model estimates, the posterior distribution for both models exhibits a complex correlation structure that is not captured in these low-dimensional summaries. Nevertheless, inspecting the posterior distribution of model estimates computed from raw model parameters can reveal important insights. Figure 1 shows each model’s joint distribution of cumulative adenoma initiation risk at ages 25 and 80. Unsurprisingly, the model that allows earlier adenoma initiation (BC-10) implies a higher cumulative risk of adenoma initiation by age 25. Despite those differences, both models have approximately the same implied cumulative adenoma initiation risk at age 80. While clinical knowledge or future evidence might favor one model over the other, this study uses both models’ posterior distributions without making further assumptions about how likely each model is.

**Figure 1:**
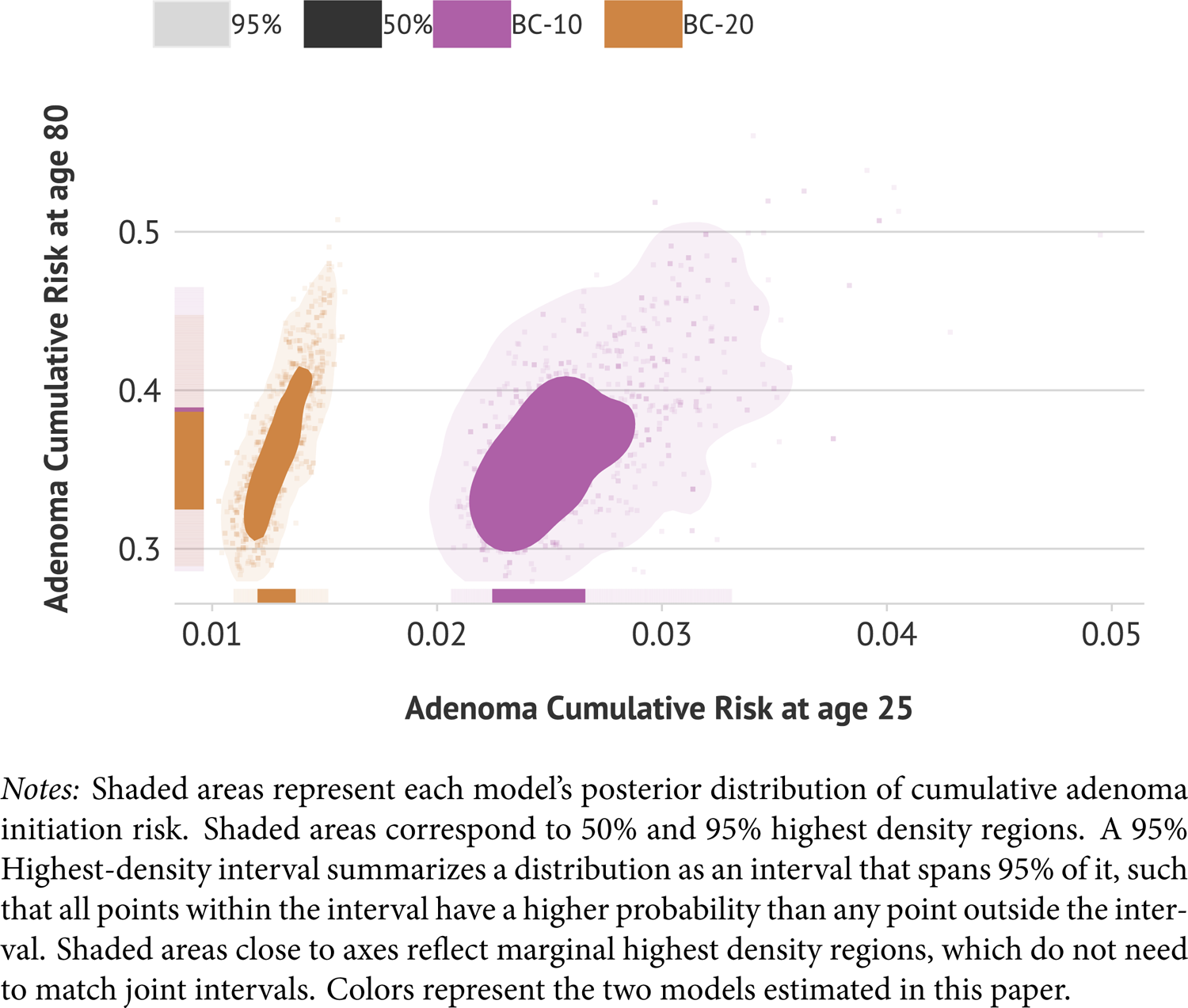
Posterior Distribution for Cumulative Adenoma Initiation Risk

Figure 2 presents results for the non-parametric birth cohort effects as incidence risk ratios. Shaded areas represent predictive credible intervals that are consistent with the data. The horizontal black line at one represents the hypothesis of no change in adenoma initiation risk by birth cohort, whereas the vertical grey line, set at 1940, represents the reference cohort against which all other cohorts are compared. This figure presents several noteworthy findings. First, it shows that the assumption of no birth cohort effects is implausible, particularly after 1940. Second, the implied relative risk ratios differ across both models (see the birth-cohort effects in table 1). This finding reinforces our suspicions that birth-cohort effects would interact with other model parameters, and estimates from one model may not translate directly to other models. Finally, these findings suggest that assuming no birth cohort effects before 1940 may be a plausible assumption.

**Figure 2:**
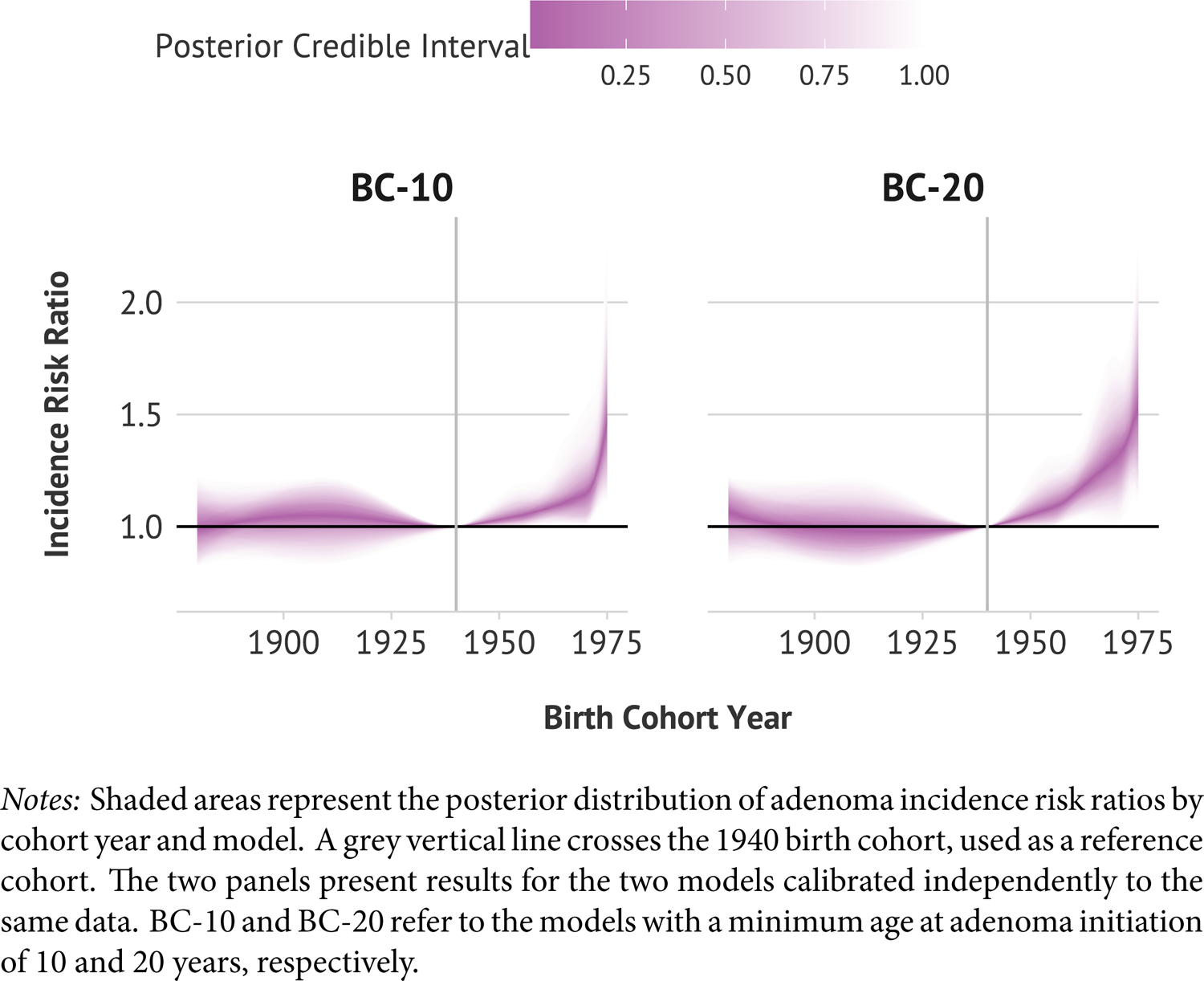
Birth Cohort Incidence Risk Ratio Estimates by Model

### 3.2 Cost-Effectiveness Estimates

Figure 3 ^4^ presents mean posterior estimates for three strategies across the eight scenarios considered in this analysis. Each scenario can be seen as one Probabilistic Sensitivity Analysis (PSA). Tables 4, 5 and 6 present the same posterior mean estimates for each combination of strategy and model, along with 95% credible intervals. Each row in figure 3 displays one model outcome measure. The top row presents Life-years gained per 1000 people. The second row shows the number of colonoscopies demanded by each screening strategy. The last column presents the incremental cost-effectiveness ratio (i.e., the number of colonoscopies one should be willing to do to gain one marginal life-year to choose that strategy). The columns represent different strategies with decreasing intensity levels from left to right. All three strategies are part of the Pareto-efficiency curve.

**Figure 3:**
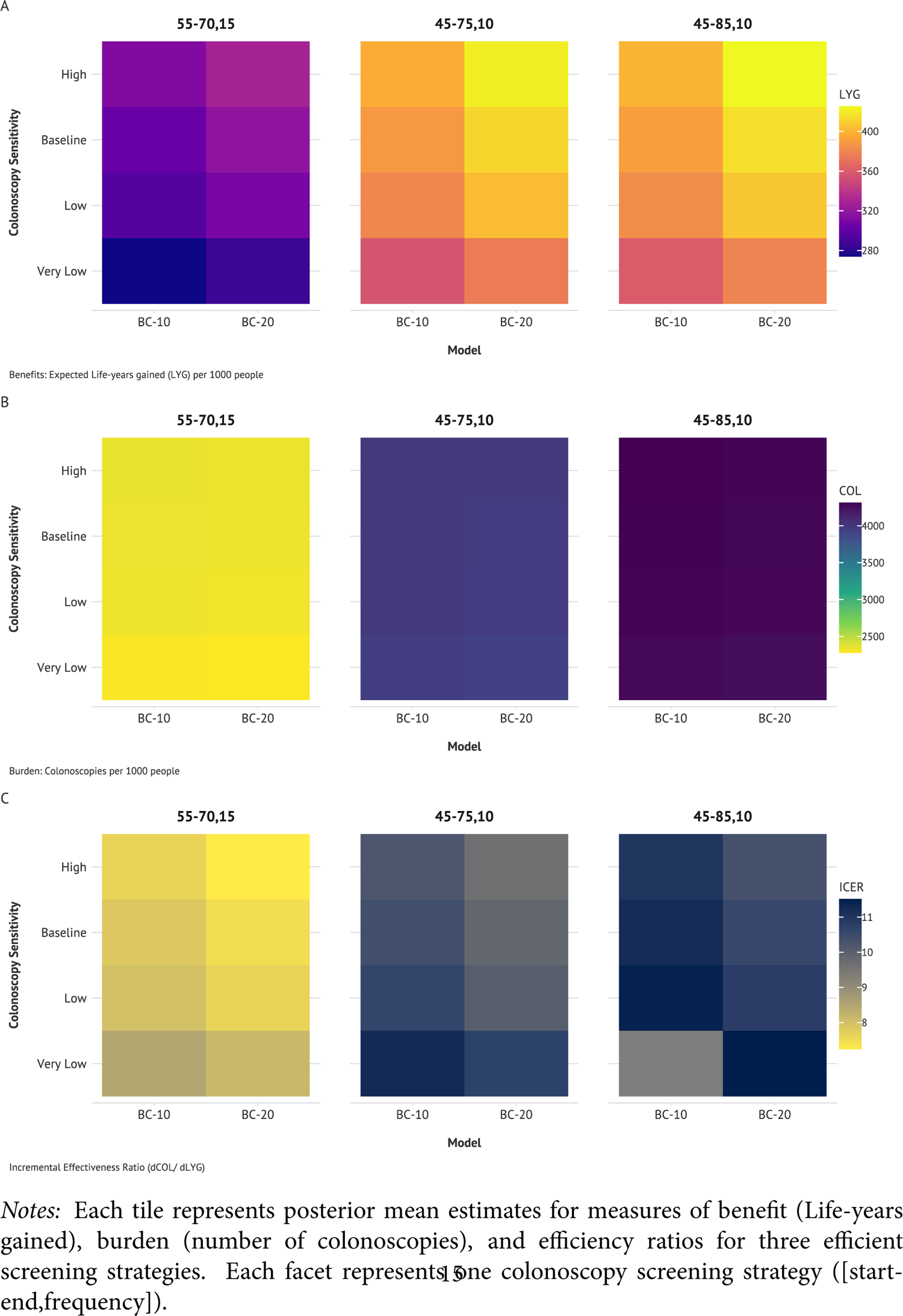
Estimates of Benefits, Burdens and Incremental Efficiency Ratios of Select CRC Colonoscopy Screening Strategies increases to 2690 (95% PI [2500, 2960]) for the 45-70,15 strategy and to 3960 (95% PI [3830, 4150]) for the 45-75,10 strategy. There are slight differences in the number of colonoscopies by sensitivity scenarios because higher sensitivity colonoscopy screening will result in a slight decrease in the required follow-up surveillance colonoscopies.

**Table 4:**
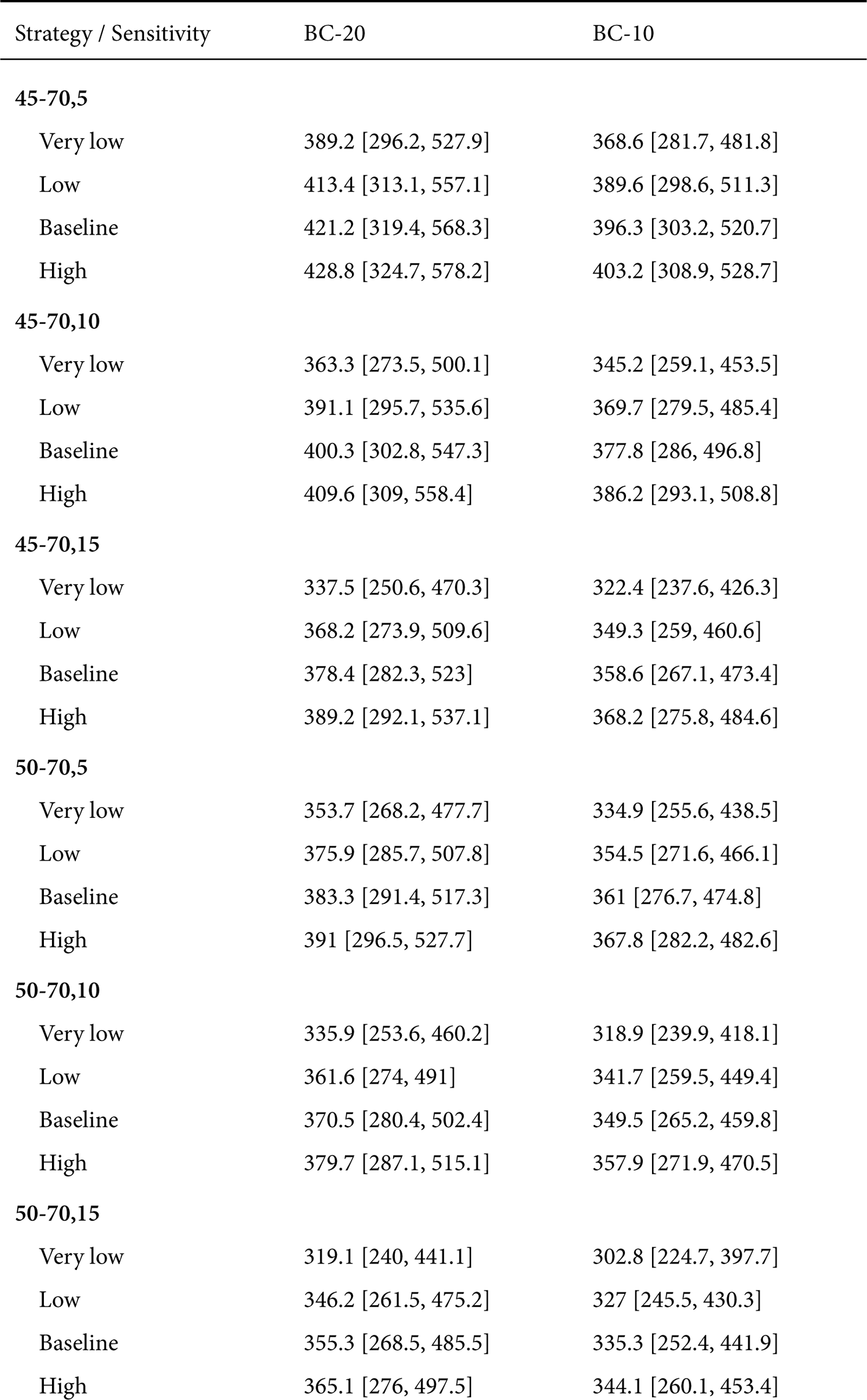
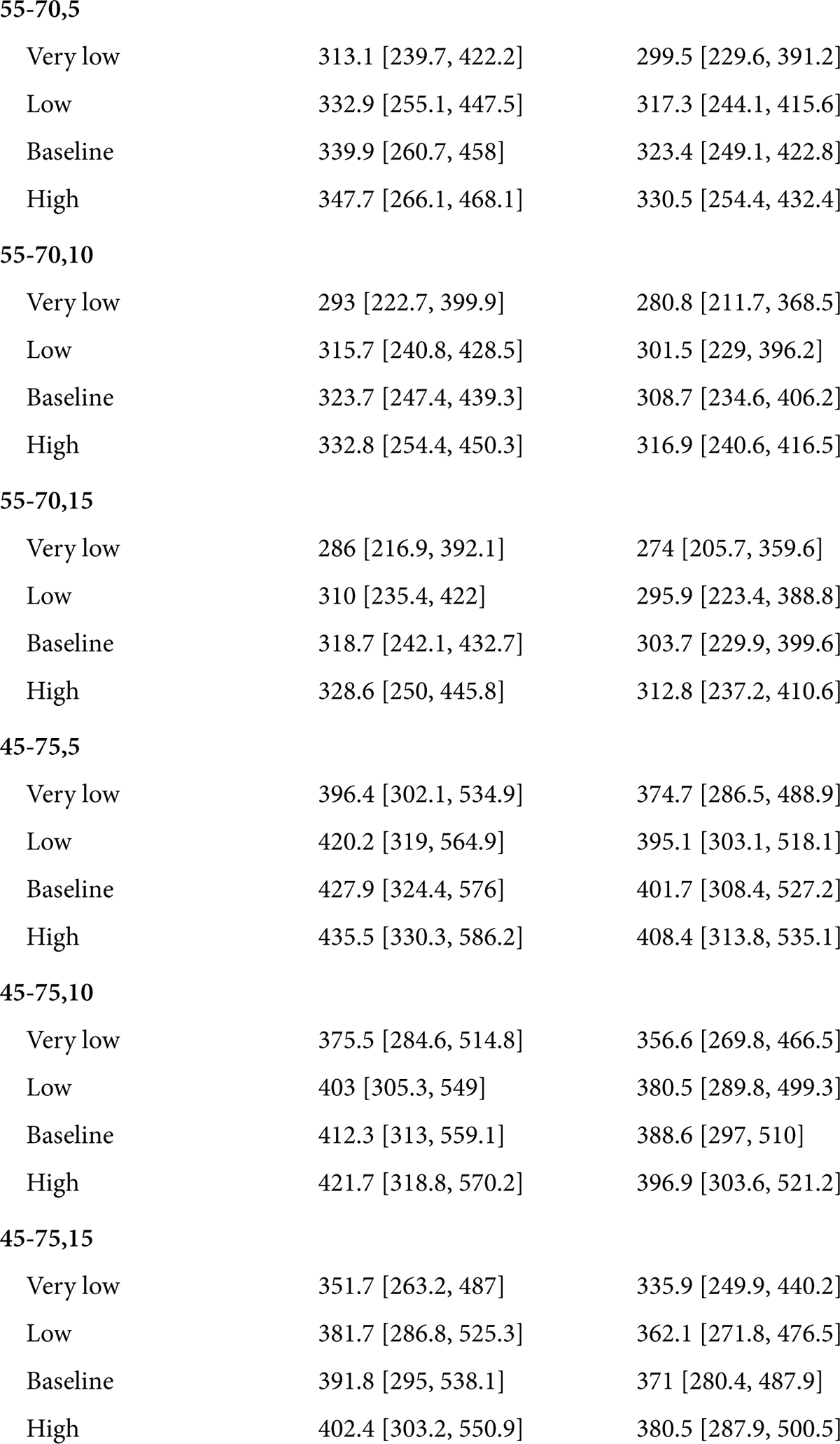
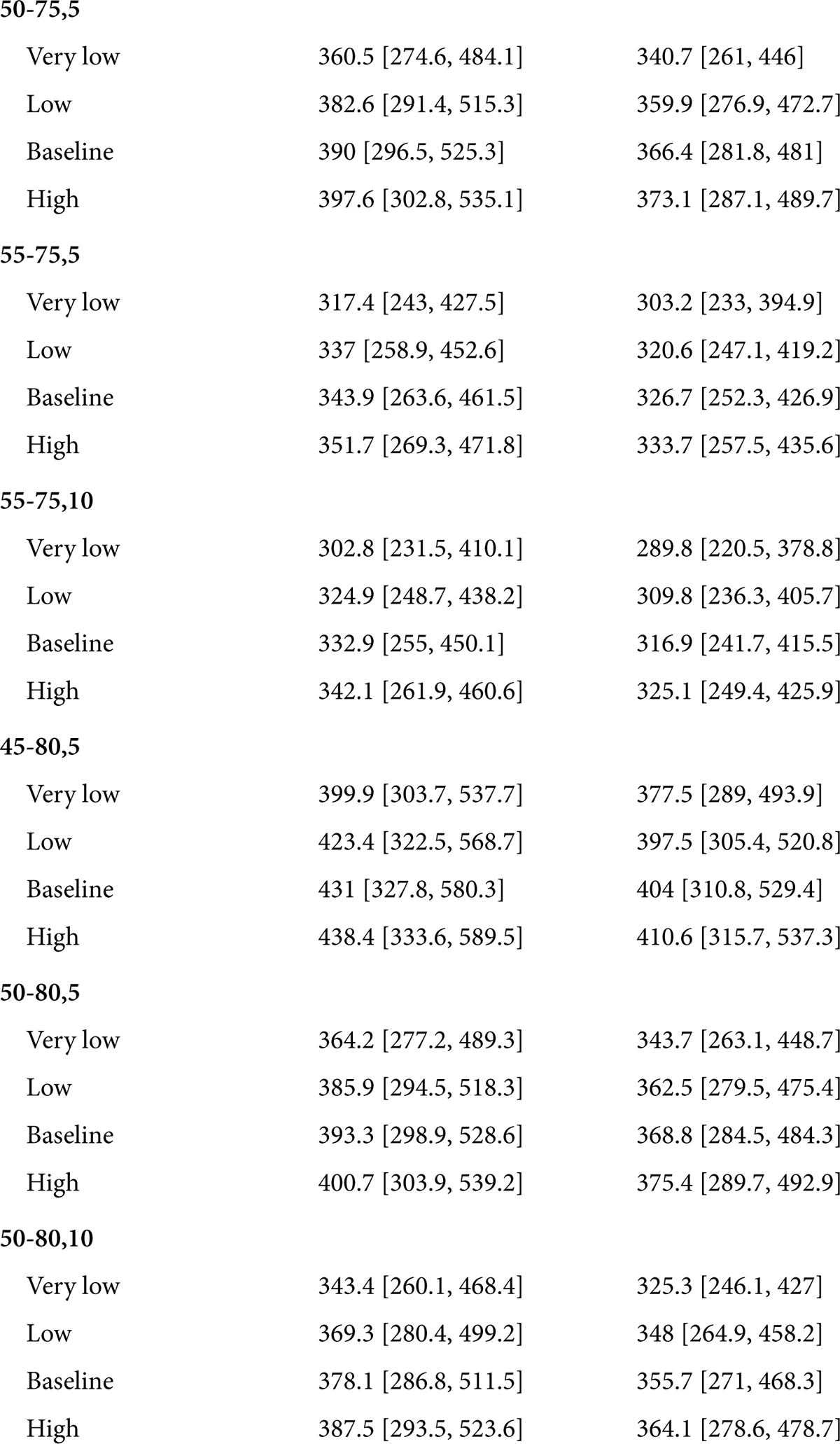
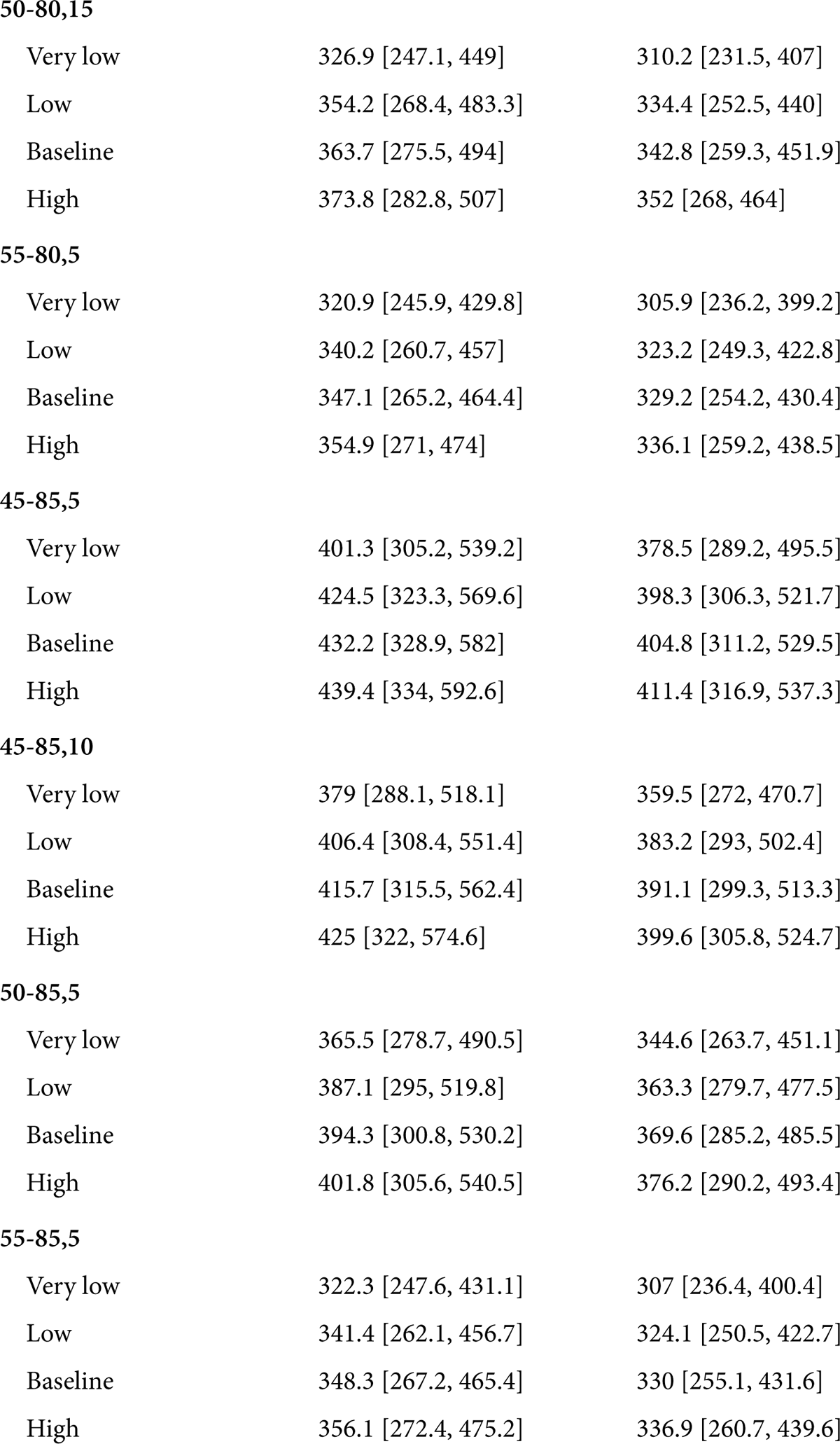
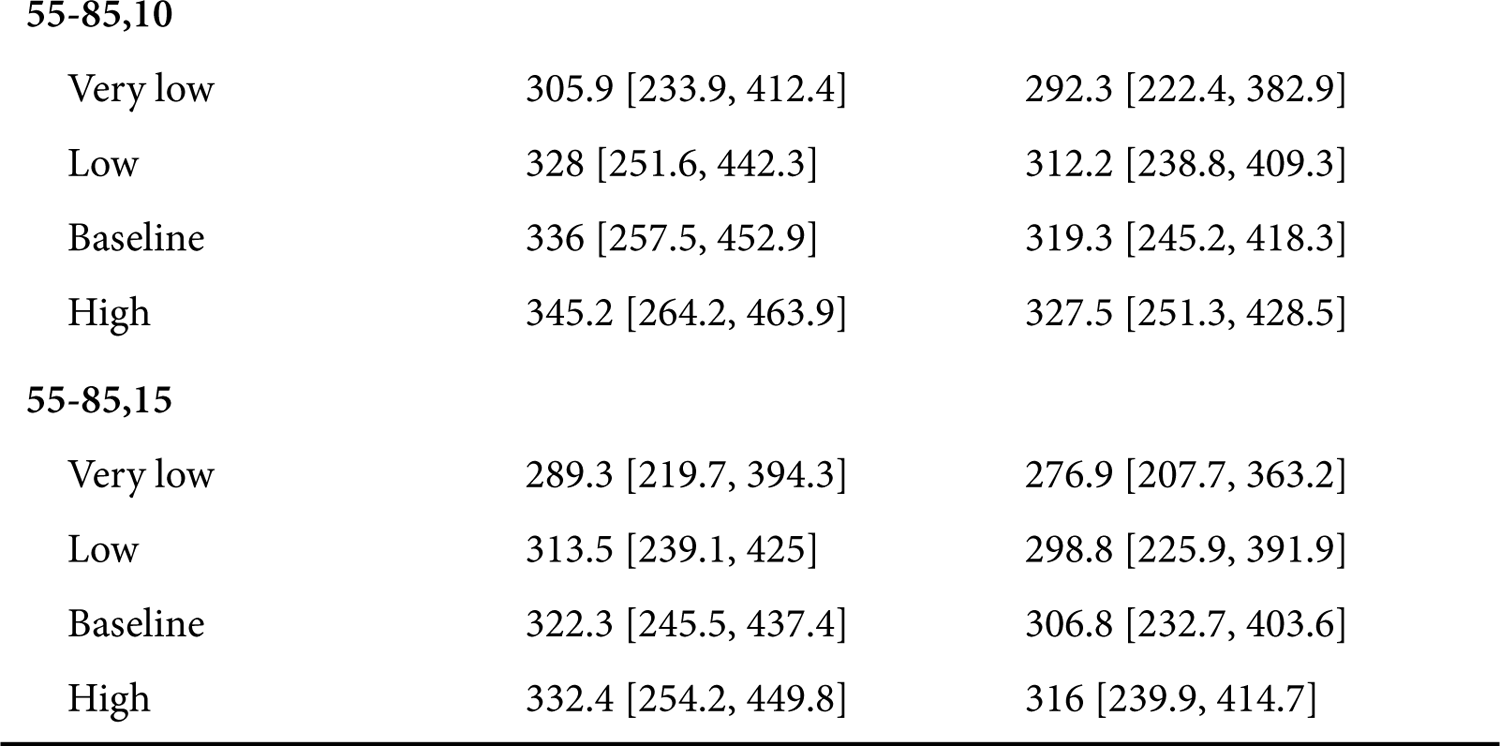
*LYG* estimates and 95 percent Credible Intervals

**Table 5:**
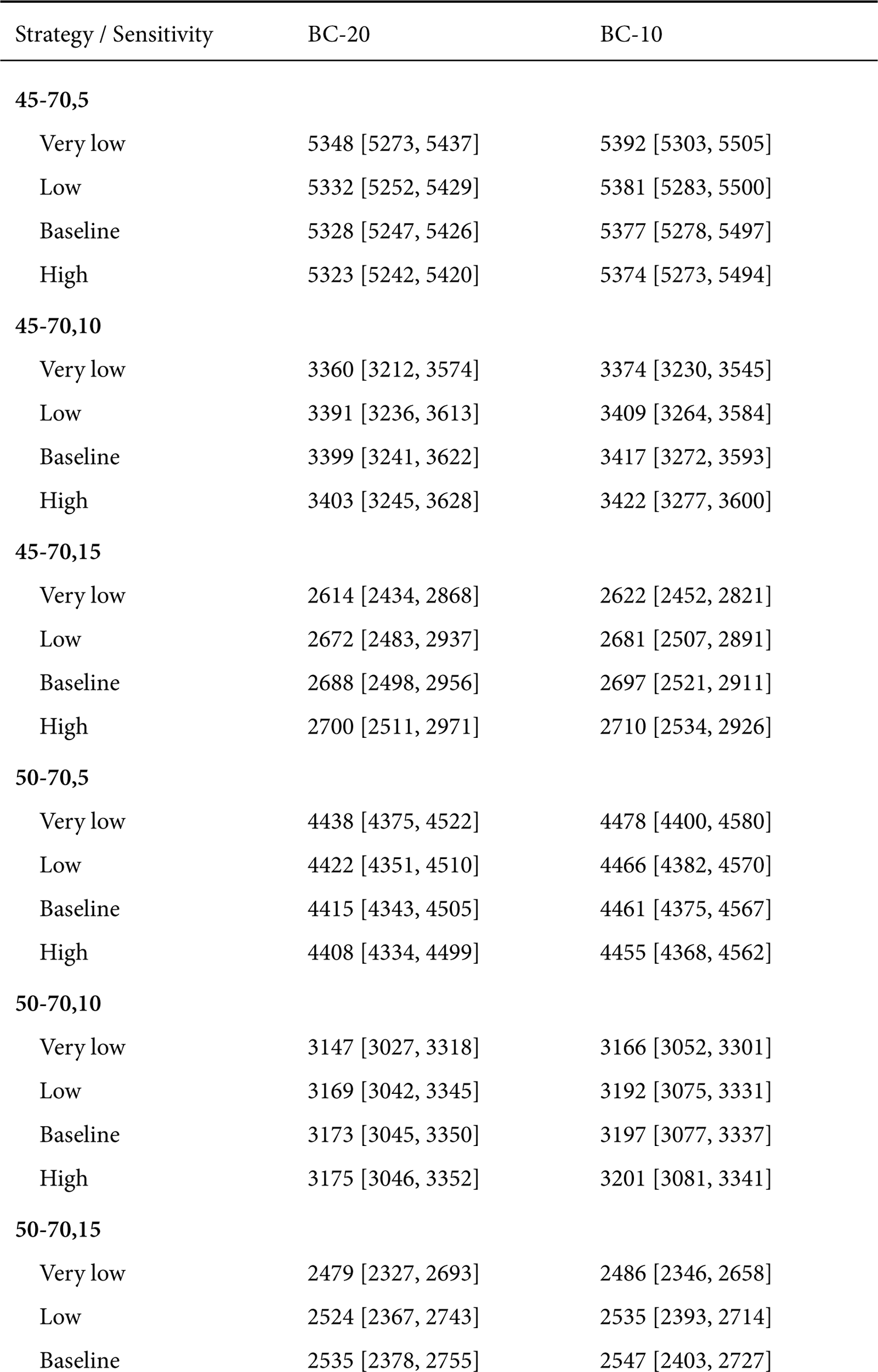
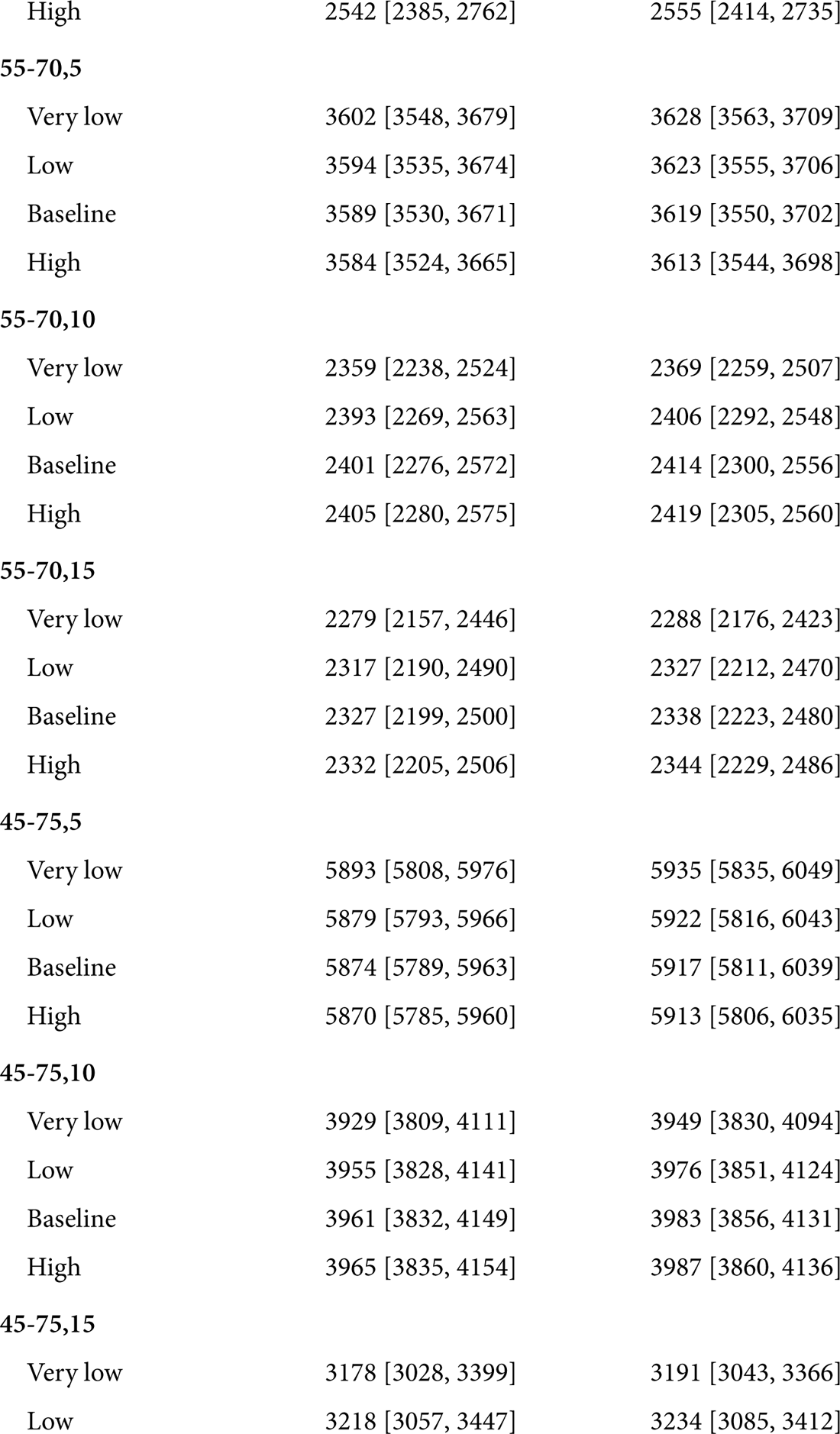
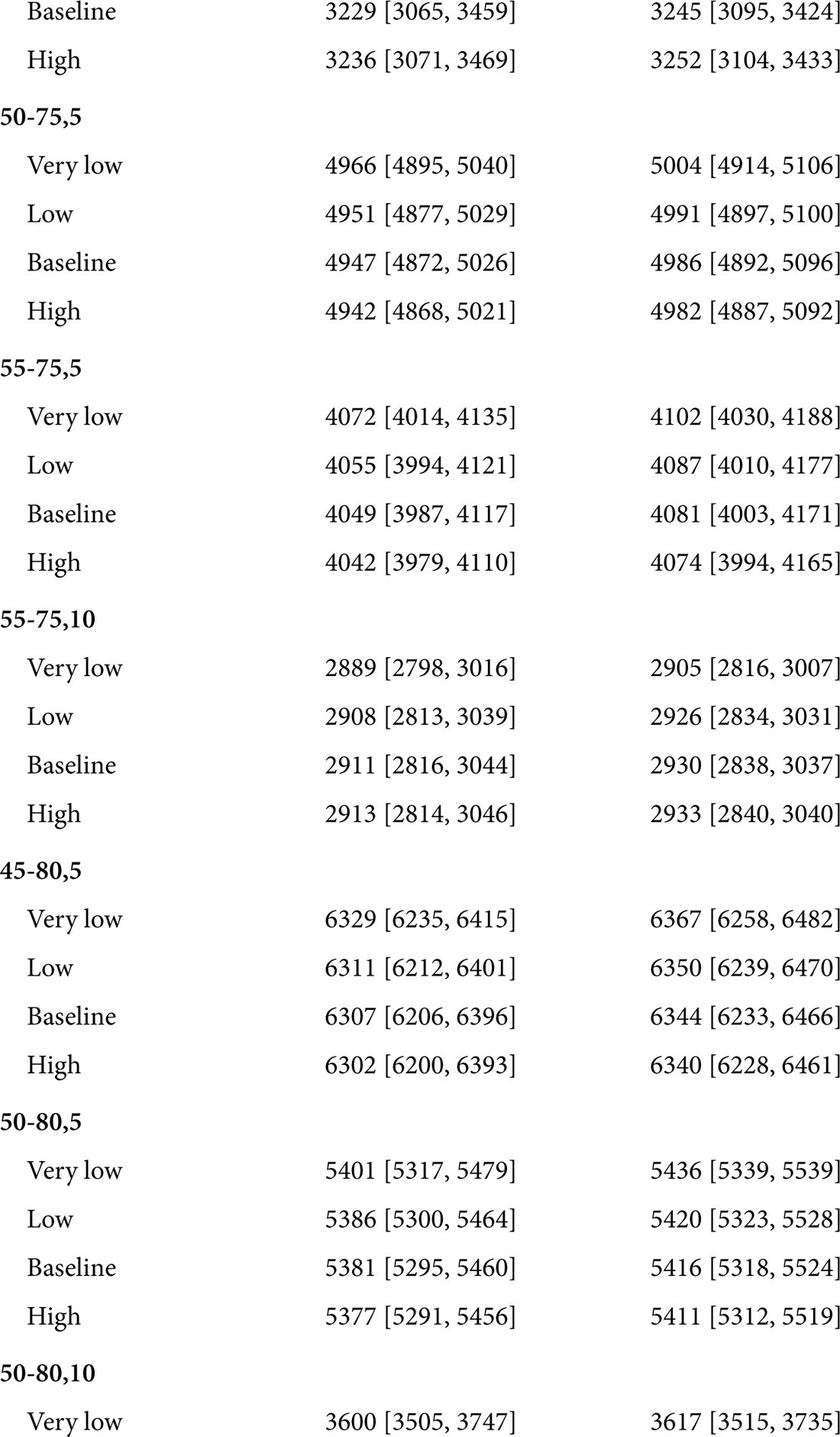
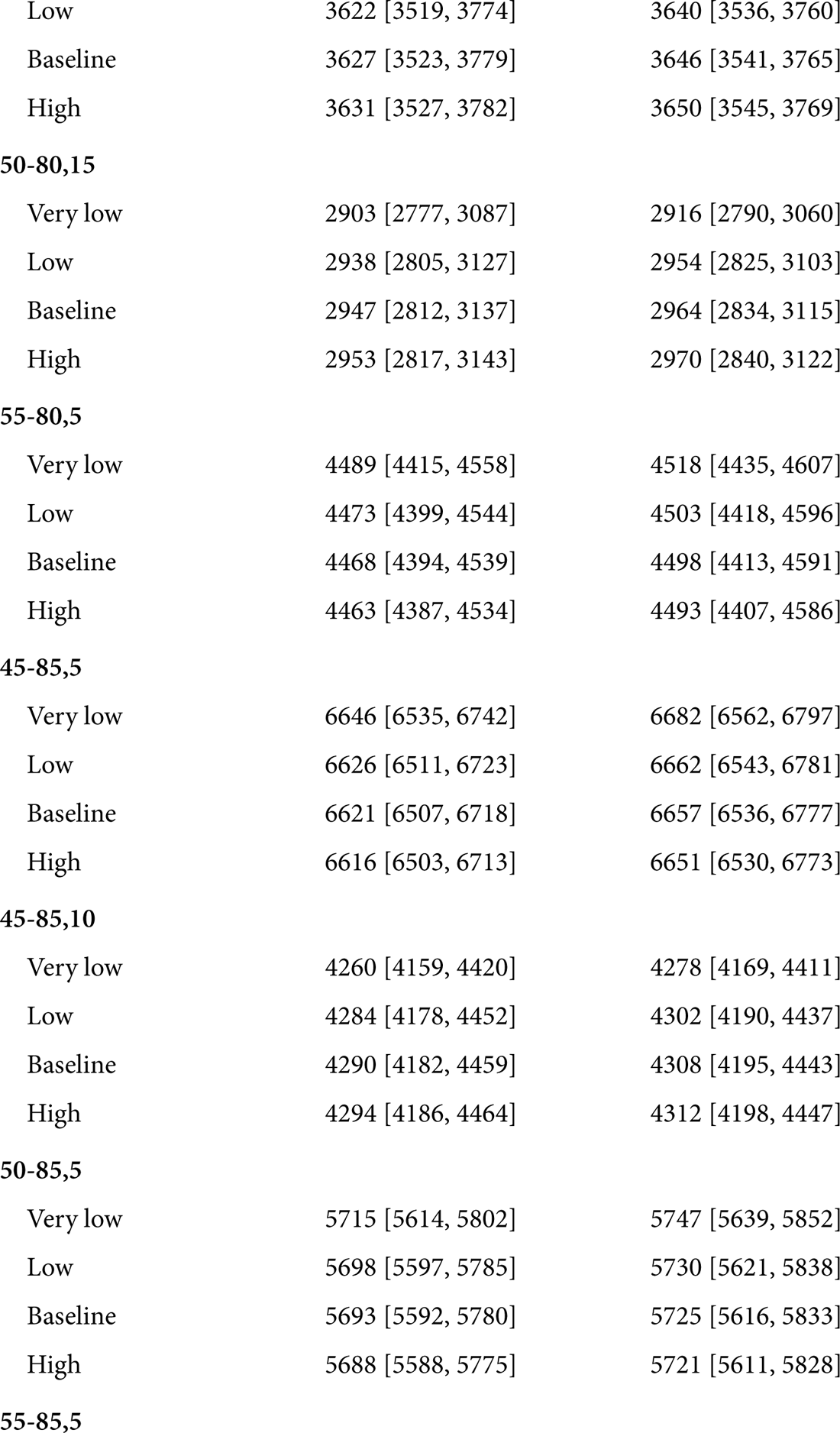
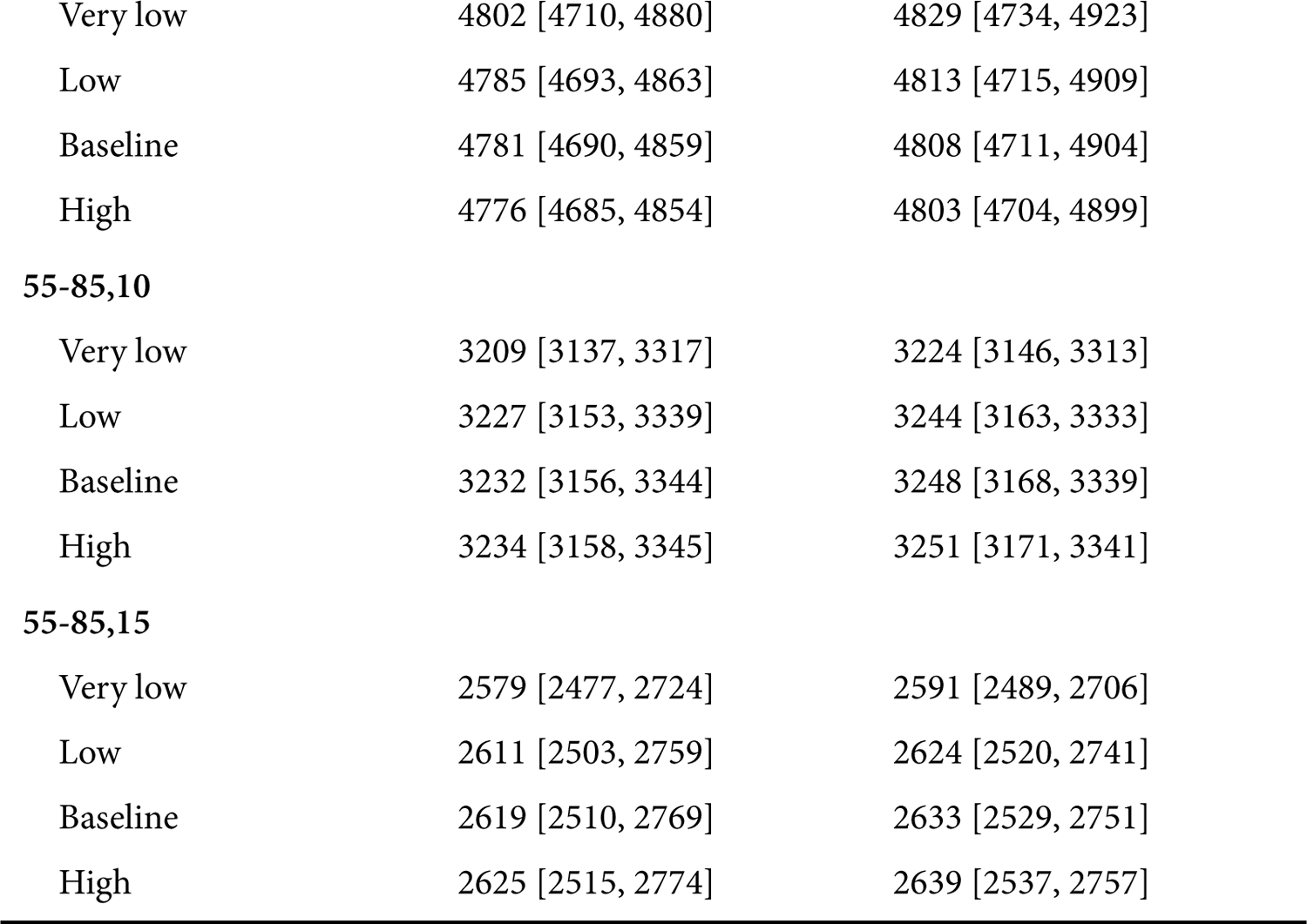
Number of Colonoscopies Estimates and 95 percent Credible Intervals

| Strategy / Sensitivity | BC-20 | BC-10 |
| --- | --- | --- |
| **45-70,5** |  |  |
| Very low | 5348 [5273, 5437] | 5392 [5303, 5505] |
| Low | 5332 [5252, 5429] | 5381 [5283, 5500] |
| Baseline | 5328 [5247, 5426] | 5377 [5278, 5497] |
| High | 5323 [5242, 5420] | 5374 [5273, 5494] |
| **45-70,10** |  |  |
| Very low | 3360 [3212, 3574] | 3374 [3230, 3545] |
| Low | 3391 [3236, 3613] | 3409 [3264, 3584] |
| Baseline | 3399 [3241, 3622] | 3417 [3272, 3593] |
| High | 3403 [3245, 3628] | 3422 [3277, 3600] |
| **45-70,15** |  |  |
| Very low | 2614 [2434, 2868] | 2622 [2452, 2821] |
| Low | 2672 [2483, 2937] | 2681 [2507, 2891] |
| Baseline | 2688 [2498, 2956] | 2697 [2521, 2911] |
| High | 2700 [2511, 2971] | 2710 [2534, 2926] |
| **50-70,5** |  |  |
| Very low | 4438 [4375, 4522] | 4478 [4400, 4580] |
| Low | 4422 [4351, 4510] | 4466 [4382, 4570] |
| Baseline | 4415 [4343, 4505] | 4461 [4375, 4567] |
| High | 4408 [4334, 4499] | 4455 [4368, 4562] |
| **50-70,10** |  |  |
| Very low | 3147 [3027, 3318] | 3166 [3052, 3301] |
| Low | 3169 [3042, 3345] | 3192 [3075, 3331] |
| Baseline | 3173 [3045, 3350] | 3197 [3077, 3337] |
| High | 3175 [3046, 3352] | 3201 [3081, 3341] |
| **50-70,15** |  |  |
| Very low | 2479 [2327, 2693] | 2486 [2346, 2658] |
| Low | 2524 [2367, 2743] | 2535 [2393, 2714] |
| Baseline | 2535 [2378, 2755] | 2547 [2403, 2727] |

Table 5: Number of Colonoscopies Estimates and 95 percent Credible Intervals (*continued*)
| Strategy / Sensitivity | BC-20 | BC-10 |
| --- | --- | --- |
| High | 2542 [2385, 2762] | 2555 [2414, 2735] |
| **55-70,5** |  |  |
| Very low | 3602 [3548, 3679] | 3628 [3563, 3709] |
| Low | 3594 [3535, 3674] | 3623 [3555, 3706] |
| Baseline | 3589 [3530, 3671] | 3619 [3550, 3702] |
| High | 3584 [3524, 3665] | 3613 [3544, 3698] |
| **55-70,10** |  |  |
| Very low | 2359 [2238, 2524] | 2369 [2259, 2507] |
| Low | 2393 [2269, 2563] | 2406 [2292, 2548] |
| Baseline | 2401 [2276, 2572] | 2414 [2300, 2556] |
| High | 2405 [2280, 2575] | 2419 [2305, 2560] |
| **55-70,15** |  |  |
| Very low | 2279 [2157, 2446] | 2288 [2176, 2423] |
| Low | 2317 [2190, 2490] | 2327 [2212, 2470] |
| Baseline | 2327 [2199, 2500] | 2338 [2223, 2480] |
| High | 2332 [2205, 2506] | 2344 [2229, 2486] |
| **45-75,5** |  |  |
| Very low | 5893 [5808, 5976] | 5935 [5835, 6049] |
| Low | 5879 [5793, 5966] | 5922 [5816, 6043] |
| Baseline | 5874 [5789, 5963] | 5917 [5811, 6039] |
| High | 5870 [5785, 5960] | 5913 [5806, 6035] |
| **45-75,10** |  |  |
| Very low | 3929 [3809, 4111] | 3949 [3830, 4094] |
| Low | 3955 [3828, 4141] | 3976 [3851, 4124] |
| Baseline | 3961 [3832, 4149] | 3983 [3856, 4131] |
| High | 3965 [3835, 4154] | 3987 [3860, 4136] |
| **45-75,15** |  |  |
| Very low | 3178 [3028, 3399] | 3191 [3043, 3366] |
| Low | 3218 [3057, 3447] | 3234 [3085, 3412] |

Table 5: Number of Colonoscopies Estimates and 95 percent Credible Intervals (*continued*)
| Strategy / Sensitivity | BC-20 | BC-10 |
| --- | --- | --- |
| Baseline | 3229 [3065, 3459] | 3245 [3095, 3424] |
| High | 3236 [3071, 3469] | 3252 [3104, 3433] |
| **50-75,5** |  |  |
| Very low | 4966 [4895, 5040] | 5004 [4914, 5106] |
| Low | 4951 [4877, 5029] | 4991 [4897, 5100] |
| Baseline | 4947 [4872, 5026] | 4986 [4892, 5096] |
| High | 4942 [4868, 5021] | 4982 [4887, 5092] |
| **55-75,5** |  |  |
| Very low | 4072 [4014, 4135] | 4102 [4030, 4188] |
| Low | 4055 [3994, 4121] | 4087 [4010, 4177] |
| Baseline | 4049 [3987, 4117] | 4081 [4003, 4171] |
| High | 4042 [3979, 4110] | 4074 [3994, 4165] |
| **55-75,10** |  |  |
| Very low | 2889 [2798, 3016] | 2905 [2816, 3007] |
| Low | 2908 [2813, 3039] | 2926 [2834, 3031] |
| Baseline | 2911 [2816, 3044] | 2930 [2838, 3037] |
| High | 2913 [2814, 3046] | 2933 [2840, 3040] |
| **45-80,5** |  |  |
| Very low | 6329 [6235, 6415] | 6367 [6258, 6482] |
| Low | 6311 [6212, 6401] | 6350 [6239, 6470] |
| Baseline | 6307 [6206, 6396] | 6344 [6233, 6466] |
| High | 6302 [6200, 6393] | 6340 [6228, 6461] |
| **50-80,5** |  |  |
| Very low | 5401 [5317, 5479] | 5436 [5339, 5539] |
| Low | 5386 [5300, 5464] | 5420 [5323, 5528] |
| Baseline | 5381 [5295, 5460] | 5416 [5318, 5524] |
| High | 5377 [5291, 5456] | 5411 [5312, 5519] |
| **50-80,10** |  |  |
| Very low | 3600 [3505, 3747] | 3617 [3515, 3735] |

Table 5: Number of Colonoscopies Estimates and 95 percent Credible Intervals (*continued*)
| Strategy / Sensitivity | BC-20 | BC-10 |
| --- | --- | --- |
| Low | 3622 [3519, 3774] | 3640 [3536, 3760] |
| Baseline | 3627 [3523, 3779] | 3646 [3541, 3765] |
| High | 3631 [3527, 3782] | 3650 [3545, 3769] |
| **50-80,15** |  |  |
| Very low | 2903 [2777, 3087] | 2916 [2790, 3060] |
| Low | 2938 [2805, 3127] | 2954 [2825, 3103] |
| Baseline | 2947 [2812, 3137] | 2964 [2834, 3115] |
| High | 2953 [2817, 3143] | 2970 [2840, 3122] |
| **55-80,5** |  |  |
| Very low | 4489 [4415, 4558] | 4518 [4435, 4607] |
| Low | 4473 [4399, 4544] | 4503 [4418, 4596] |
| Baseline | 4468 [4394, 4539] | 4498 [4413, 4591] |
| High | 4463 [4387, 4534] | 4493 [4407, 4586] |
| **45-85,5** |  |  |
| Very low | 6646 [6535, 6742] | 6682 [6562, 6797] |
| Low | 6626 [6511, 6723] | 6662 [6543, 6781] |
| Baseline | 6621 [6507, 6718] | 6657 [6536, 6777] |
| High | 6616 [6503, 6713] | 6651 [6530, 6773] |
| **45-85,10** |  |  |
| Very low | 4260 [4159, 4420] | 4278 [4169, 4411] |
| Low | 4284 [4178, 4452] | 4302 [4190, 4437] |
| Baseline | 4290 [4182, 4459] | 4308 [4195, 4443] |
| High | 4294 [4186, 4464] | 4312 [4198, 4447] |
| **50-85,5** |  |  |
| Very low | 5715 [5614, 5802] | 5747 [5639, 5852] |
| Low | 5698 [5597, 5785] | 5730 [5621, 5838] |
| Baseline | 5693 [5592, 5780] | 5725 [5616, 5833] |
| High | 5688 [5588, 5775] | 5721 [5611, 5828] |
| **55-85,5** |  |  |

Table 5: Number of Colonoscopies Estimates and 95 percent Credible Intervals (*continued*)
| Strategy / Sensitivity | BC-20 | BC-10 |
| --- | --- | --- |
| Very low | 4802 [4710, 4880] | 4829 [4734, 4923] |
| Low | 4785 [4693, 4863] | 4813 [4715, 4909] |
| Baseline | 4781 [4690, 4859] | 4808 [4711, 4904] |
| High | 4776 [4685, 4854] | 4803 [4704, 4899] |
| **55-85,10** |  |  |
| Very low | 3209 [3137, 3317] | 3224 [3146, 3313] |
| Low | 3227 [3153, 3339] | 3244 [3163, 3333] |
| Baseline | 3232 [3156, 3344] | 3248 [3168, 3339] |
| High | 3234 [3158, 3345] | 3251 [3171, 3341] |
| **55-85,15** |  |  |
| Very low | 2579 [2477, 2724] | 2591 [2489, 2706] |
| Low | 2611 [2503, 2759] | 2624 [2520, 2741] |
| Baseline | 2619 [2510, 2769] | 2633 [2529, 2751] |
| High | 2625 [2515, 2774] | 2639 [2537, 2757] |

**Table 6:** Efficiency Ratio Estimates and 95 percent Credible Intervals

| Strategy / Sensitivity | BC-20 | BC-10 |
| --- | --- | --- |
| **45-70,5** |  |  |
| Very low | Dominated | Dominated |
| Low | Dominated | Dominated |
| Baseline | Dominated | Dominated |
| High | Dominated | Dominated |
| **45-70,10** |  |  |
| Very low | 9.464 [7.089, 11.94] | 9.942 [7.683, 12.51] |
| Low | 8.872 [6.643, 11.19] | 9.375 [7.27, 11.72] |
| Baseline | 8.684 [6.508, 10.92] | 9.194 [7.146, 11.48] |
| High | 8.498 [6.379, 10.7] | 9.006 [7.035, 11.27] |
| **45-70,15** |  |  |
| Very low | 7.922 [6.021, 9.86] | 8.271 [6.485, 10.33] |
| Low | 7.419 [5.661, 9.245] | 7.8 [6.139, 9.68] |
| Baseline | 7.257 [5.542, 9.042] | 7.642 [6.026, 9.446] |
| High | 7.088 [5.427, 8.816] | 7.475 [5.917, 9.221] |
| **50-70,5** |  |  |
| Very low | Dominated | Dominated |
| Low | Dominated | Dominated |
| Baseline | Dominated | Dominated |
| High | Dominated | Dominated |
| **50-70,10** |  |  |
| Very low | Dominated | Dominated |
| Low | Dominated | Dominated |
| Baseline | Dominated | Dominated |
| High | Dominated | Dominated |
| **50-70,15** |  |  |
| Very low | 7.939 [6.028, 9.921] | 8.348 [6.499, 10.43] |
| Low | 7.449 [5.705, 9.248] | Dominated |
| Baseline | 7.287 [5.58, 9.054] | Dominated |
| High | 7.11 [5.44, 8.853] | Dominated |

Table 6: Efficiency Ratio Estimates and 95 percent Credible Intervals (*continued*)
| Strategy / Sensitivity | BC-20 | BC-10 |
| --- | --- | --- |
| **55-70,5** |  |  |
| Very low | Dominated | Dominated |
| Low | Dominated | Dominated |
| Baseline | Dominated | Dominated |
| High | Dominated | Dominated |
| **55-70,10** |  |  |
| Very low | Dominated | Dominated |
| Low | Dominated | Dominated |
| Baseline | Dominated | Dominated |
| High | Dominated | Dominated |
| **55-70,15** |  |  |
| Very low | 8.139 [6.193, 10.1] | 8.487 [6.604, 10.57] |
| Low | 7.633 [5.825, 9.444] | 7.991 [6.256, 9.911] |
| Baseline | 7.454 [5.684, 9.242] | 7.821 [6.128, 9.661] |
| High | 7.246 [5.525, 9.001] | 7.614 [5.988, 9.437] |
| **45-75,5** |  |  |
| Very low | 15.24 [11.07, 19.62] | 16.13 [12.02, 20.78] |
| Low | 14.34 [10.39, 18.53] | 15.26 [11.38, 19.59] |
| Baseline | 14.07 [10.17, 18.14] | 15 [11.18, 19.28] |
| High | Dominated | 14.74 [11.01, 18.93] |
| **45-75,10** |  |  |
| Very low | 10.71 [7.945, 13.58] | 11.27 [8.638, 14.29] |
| Low | 10.05 [7.417, 12.74] | 10.63 [8.18, 13.36] |
| Baseline | 9.833 [7.266, 12.45] | 10.42 [8.037, 13.13] |
| High | 9.622 [7.128, 12.19] | 10.21 [7.917, 12.83] |
| **45-75,15** |  |  |
| Very low | Dominated | Dominated |
| Low | Dominated | Dominated |
| Baseline | Dominated | Dominated |

Table 6: Efficiency Ratio Estimates and 95 percent Credible Intervals (*continued*)
| Strategy / Sensitivity | BC-20 | BC-10 |
| --- | --- | --- |
| High | Dominated | Dominated |
| **50-75,5** |  |  |
| Very low | Dominated | Dominated |
| Low | Dominated | Dominated |
| Baseline | Dominated | Dominated |
| High | Dominated | Dominated |
| **55-75,5** |  |  |
| Very low | Dominated | Dominated |
| Low | Dominated | Dominated |
| Baseline | Dominated | Dominated |
| High | Dominated | Dominated |
| **55-75,10** |  |  |
| Very low | Dominated | Dominated |
| Low | Dominated | Dominated |
| Baseline | Dominated | Dominated |
| High | Dominated | Dominated |
| **45-80,5** |  |  |
| Very low | 16.23 [11.73, 21.03] | 17.18 [12.75, 22.09] |
| Low | 15.29 [11.03, 19.71] | 16.27 [12.08, 20.93] |
| Baseline | 15.01 [10.8, 19.37] | 15.99 [11.9, 20.56] |
| High | 14.74 [10.63, 19.02] | 15.72 [11.7, 20.23] |
| **50-80,5** |  |  |
| Very low | Dominated | Dominated |
| Low | Dominated | Dominated |
| Baseline | Dominated | Dominated |
| High | Dominated | Dominated |
| **50-80,10** |  |  |
| Very low | Dominated | Dominated |
| Low | Dominated | Dominated |

Table 6: Efficiency Ratio Estimates and 95 percent Credible Intervals (*continued*)
| Strategy / Sensitivity | BC-20 | BC-10 |
| --- | --- | --- |
| Baseline | Dominated | Dominated |
| High | Dominated | Dominated |
| **50-80,15** |  |  |
| Very low | Dominated | Dominated |
| Low | Dominated | Dominated |
| Baseline | Dominated | Dominated |
| High | Dominated | Dominated |
| **55-80,5** |  |  |
| Very low | Dominated | Dominated |
| Low | Dominated | Dominated |
| Baseline | Dominated | Dominated |
| High | Dominated | Dominated |
| **45-85,5** |  |  |
| Very low | 16.98 [12.25, 21.99] | 17.98 [13.29, 23.22] |
| Low | 16.01 [11.53, 20.7] | 17.03 [12.6, 21.92] |
| Baseline | 15.72 [11.29, 20.33] | 16.75 [12.41, 21.53] |
| High | 15.44 [11.08, 19.98] | 16.47 [12.23, 21.2] |
| **45-85,10** |  |  |
| Very low | 11.51 [8.498, 14.64] | Dominated |
| Low | 10.79 [7.931, 13.71] | 11.42 [8.74, 14.47] |
| Baseline | 10.57 [7.759, 13.44] | 11.2 [8.576, 14.17] |
| High | 10.34 [7.602, 13.13] | 10.98 [8.43, 13.86] |
| **50-85,5** |  |  |
| Very low | Dominated | Dominated |
| Low | Dominated | Dominated |
| Baseline | Dominated | Dominated |
| High | Dominated | Dominated |
| **55-85,5** |  |  |
| Very low | Dominated | Dominated |

Table 6: Efficiency Ratio Estimates and 95 percent Credible Intervals (*continued*)
| Strategy / Sensitivity | BC-20 | BC-10 |
| --- | --- | --- |
| Low | Dominated | Dominated |
| Baseline | Dominated | Dominated |
| High | Dominated | Dominated |
| **55-85,10** |  |  |
| Very low | Dominated | Dominated |
| Low | Dominated | Dominated |
| Baseline | Dominated | Dominated |
| High | Dominated | Dominated |
| **55-85,15** |  |  |
| Very low | Dominated | Dominated |
| Low | Dominated | Dominated |
| Baseline | Dominated | Dominated |
| High | Dominated | Dominated |

**Table 7:** Back-to-back Colonoscopy Studies

| Adenoma Size | Study | $n_1$ | $n_2$ | AMR |
| --- | --- | --- | --- | --- |
| <=5 mm | Hixon, 1991 | 89 | 17 | 16.04 |
| <=5 mm | Rex, 1997 | 217 | 81 | 27.18 |
| <=5 mm | Rex, 2003 I | 69 | 23 | 25.00 |
| <=5 mm | Rex, 2003 II | 12 | 8 | 40.00 |
| <=5 mm | Harrison, 2004 | 49 | 22 | 30.99 |
| 6-9 mm | Hixon, 1991 | 50 | 7 | 12.28 |
| 6-9 mm | Rex, 1997 | 42 | 6 | 12.50 |
| 6-9 mm | Rex, 2003 I | 8 | 0 | 0.00 |
| 6-9 mm | Rex, 2003 II | 3 | 2 | 40.00 |
| 6-9 mm | Harrison, 2004 | 5 | 1 | 16.67 |
| >= 10 mm | Hixon, 1991 | 58 | 0 | 0.00 |
| >= 10 mm | Rex, 1997 | 30 | 2 | 6.25 |
| >= 10 mm | Rex, 2003 I | 2 | 0 | 0.00 |
| >= 10 mm | Rex, 2003 II | 1 | 0 | 0.00 |
| >= 10 mm | Harrison, 2004 | 3 | 0 | 0.00 |

Figure 3 A shows that *LYG* estimates are highly contingent on sensitivity assumptions. Under baseline sensitivity and model assumptions (Sensitivity R Baseline and Model R BC-20), Strategy 45-75,10 results in 412 (95% PI [313, 559]) *LYG* / 1000 people. Under a high sensitivity scenario, the same strategy results in 422 (95% PI [319, 570]) *LYG* / 1000 people. The benefit from the same policy decreases to 375 (95% PI [285, 515]) if sensitivity is very low - a 12% difference in effectiveness.

Figure 3 B demonstrates that the number of colonoscopies performed is similar across sensitivity scenarios and natural history assumptions and highly contingent on the intensity of the screening strategy. Under baseline assumptions, strategy 55-70,15 is expected to result in 2330 (95% PI [2200, 2500]) colonoscopies per 1000 individuals. This estimate II. Similarly, the minimum age to start adenoma could also be expressed as a continuous variable and be regarded as one natural history parameter, but we chose two specific values to facilitate model calibration and comparisons. Therefore, the scenarios chosen in this figure represent the boundary cells of a hypothetical fine-grained design that one could run with an unlimited computing budget.

Figure 3 C shows how the cost-effectiveness ratios change across assumptions. As expected, lower sensitivity assumptions imply higher efficiency ratios. This result demonstrates the robustness of colonoscopy screening strategies. Even under the most pessimistic combination of scenario and model specification, the efficiency ratio estimate for strategy 45-75,10 is 11.3 (95% PI [8.64, 14.3]). Under baseline assumptions, the efficiency ratio estimates improves to 9.83 (95% PI [7.27, 12.4]) colonoscopies per life years gained (on the margin).

### 3.3 Cost-Effectiveness Efficiency Frontier

Figure 4 presents the set of non-dominated screening strategies at the cost-effectiveness efficiency plane, with the burden (number of colonoscopies per 1000 people) in the horizontal axis and benefits (life-years gained per 1000 people) in the vertical axis. 95 % prediction intervals for *LYG* are displayed as shaded areas. The figure demonstrates that estimates differ by model and sensitivity, but the efficiency frontier is reasonably stable across model specifications and scenarios. Most strategies considered efficient under one scenario are also efficient in other scenarios. Table 4 presents efficiency ratio estimates for each non-dominated policy. Figure 4 also shows that the differences between specifications BC-10 and BC-20 and relatively small when contrasted with the uncertainty resulting from the posterior distribution of natural history parameters. There is a wide range of possible *LYG* outcomes for each given policy.

**Figure 4:**
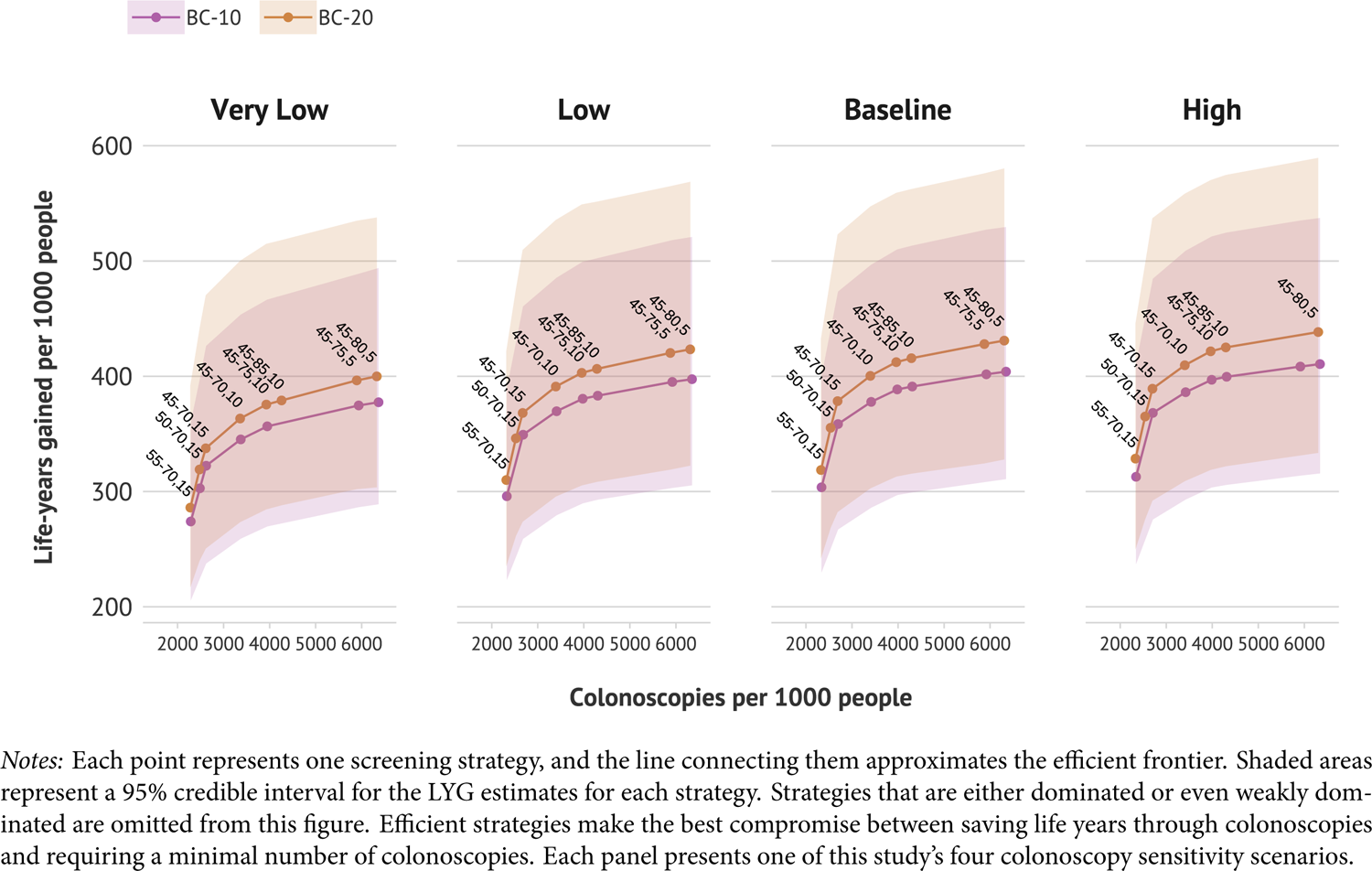
Cost-Effectiveness of Screening Strategies by Sensitivity and Model

## 4 Discussion

To our knowledge, this is the first CRC microsimulation analysis to produce and use a posterior distribution of birth-cohort effects in adenoma initiation risk. The birth-cohort model used in this study offers one alternative to explain the increase in CRC incidence. This approach mirrors the prior approach used by CISNET models (Knudsen et al. 2021a) but produces a posterior distribution for birth cohort effect parameters. This study also estimated birth cohort effects for two alternative CRC-SPIN specifications and demonstrated that those estimates are contingent on other model assumptions, such as the minimum age at adenoma initiation.

This study’s estimates of CRC screening cost-effectiveness measures varied substantially across and within scenarios. These results are in line with previously published cost-effectiveness analyses. For instance, life-years gained estimates for strategy 45-75,10 was 412 (95% PI [313, 559]) *LYG* / 1000 people for the specification BC-20 and 389 (95% PI [297, 510]) for specification BC-10. The range of *LYG* for the same strategies presented by Knudsen et al. (2021a) across the three CISNET models was 291-361. The point estimate from our study falls within the range of outcomes presented in Knudsen et al. (2021a), and the range of outcomes in our study has a wide overlap with ranges presented previously. Our estimate’s upper bound is somewhat higher than the baseline scenarios. These results reflect uncertainty in the natural history of parameters and birth-cohort effects that are not incorporated in the baseline scenario used in Knudsen et al. (2021a). These results demonstrate that natural history parameter uncertainty can be as relevant as model specification differences. As computational resources become more widely available and more microsimulation models transition to performing PSAs, the uncertainty intervals reported in multi-model analyses will better characterize the full range of uncertainty implied by different models.

Our findings highlight the importance of access to high-sensitivity screening to ensure that the benefits of screening are equitably distributed in the population. If access to high-sensitivity screening is unequal, then the benefits of screening will not be equally distributed in the population. For instance, a person who receives decennial colonoscopy screening from 45 to 75 years with very low colonoscopy sensitivity is estimated to have a 12% decrease in *LYG* compared to another person who receives the same screening strategy but with high-sensitivity colonoscopy.

Despite the uncertainties associated with *LYG* estimates, this study projected that currently-recommended colonoscopy screening strategies would be cost-effective across many conditions. While the cost-effectiveness of screening strategies changed across scenarios, the cost-effectiveness frontier remained stable across scenarios.

This study presents a series of limitations and opportunities for further extensions. First, we only present one explanation for increased CRC risk through an overall increase in the risk of adenoma initiation based on birth cohorts. This model does not include alternative model specifications that are plausible. For instance, the increased risk of CRC could be concentrated in specific locations of the large intestine. Moreover, birth cohort effects might also be present in other model parameters, including those that control growth. These alternative hypotheses represent alternative biological explanations for increased CRC incidence, and those differences can be decision-relevant. Future investigations can expand the space of plausible models and further investigate the robustness of screening strategies across those additional models that have yet to be proposed.

Future work can also build upon the eight probabilistic sensitivity analyses performed in this analysis and further investigate the significance of uncertainty by computing the opportunity cost associated with sub-optimal screening strategies and performing Value of Information (Claxton and Sculpher 2006) analyses. Because the efficiency ratio intervals of currently-recommended strategies are relatively low, those analyses are not expected to change existing recommendations. Nevertheless, one can still calculate the expected opportunity loss of each strategy on willingness to pay thresholds to determine *the* optimal strategy given a specified willingness to pay threshold using a net monetary benefit (NMB) measure. We refrain from doing so in this analysis for a few reasons. First, existing cost-effectiveness analyses of CRC screening strategies do not use QALYs as the outcome measure and do not consider the costs of colonoscopies in dollars, nor do they estimate all burdens of colonoscopy screening using a dollar amount. Second, the purpose of this paper was to compare our results to those previously published in Knudsen et al. (2021a) directly, and changing the analytical framework and measures (i.e., changing the outcome from *LYG* to discounted QALYs) would work against this goal. Either way, this study removes barriers to adopting Value of Information analyses with microsimulation models, which can still be performed based on our results.

## 5 Conclusion

This paper calibrated two specifications of the CRCSPIN model, including birth-cohort effects on adenoma initiation, and presented a stress test of CRC colonoscopy screening strategies. This study reported prediction intervals for benefits, burden, and incremental efficiency ratio for all CRC screening strategies considered in US screening guidelines across four colonoscopy sensitivity scenarios and two CRC-SPIN model specifications. Our results demonstrate that current USPSTF recommendations are robust under a wide range of conditions. In particular, decennial colonoscopy screening from 45 to 75 years was projected to remain at the cost-effectiveness frontier regardless of model specification or sensitivity level.

## Data Availability

This is a computational study based on CRCSPIN, a model part of the Cancer Intervention and Surveillance Modeling Network (CISNET). Documentation of CISNET models used to produuce the results presented in this study can be found at https://cisnet.cancer.gov/colorectal/profiles.html. Interested researchers can contact authors directly for more insight into the CISNET models.

## Acknowledgements

This research was supported by a Rothenberg Dissertation Award provided by the Pardee RAND Graduate School and by the National Cancer Institute (NCI) as part of the Cancer Intervention and Surveillance Modeling Network (CISNET) through grant U01-CA253913. This research used resources of the Argonne Leadership Computing Facility, a DOE Office of Science User Facility supported under Contract DE-AC02-06CH11357. I thank the Argonne Leadership Computing Facility staff for their timely and critical support. This research was completed with resources provided by the Laboratory Computing Resource Center at Argonne National Laboratory. The content is solely the author’s responsibility and does not necessarily represent the views of any sponsor.

## 7 Appendix I: Model Specification

### 7.1 Adenoma Growth

Adenoma growth is simulated as follows. For each lesion, CRC-SPIN simulates the lesion time to reach 10 mm (t_10ij_) using a Fréchet (inverse Weibull) distribution, with cumulative distribution function (2). This function is defined separately for lesions in the colon and in the rectum, which results in four parameters (β_1t_, β_2t_, β_1r_, β_2r_). CRC-SPIN allows variability in growth rates by sampling *t*_10ij_ from the cdf given by eq. (2).

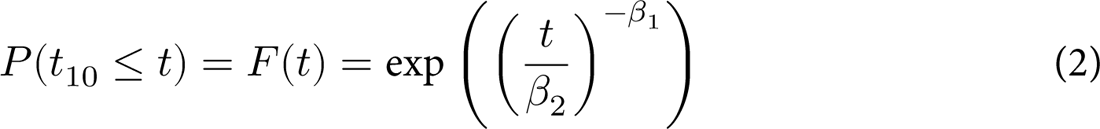

Eq. (3) describes the size (diameter) of each adenoma at any point in time *d*_ij_(t). This equation is a Richard’s Growth model, following a *W*_0_-form (Tjørve and Tjørve 2010). *d*_∞_ R 50 mm is the maximum adenoma size, *d*_0_ = 1 mm is the size at adenoma initiation, r_ij_ is the maximum adenoma growth rate, *t* is time after adenoma initiation, and *p* determines the relative size at maximum growth rate.

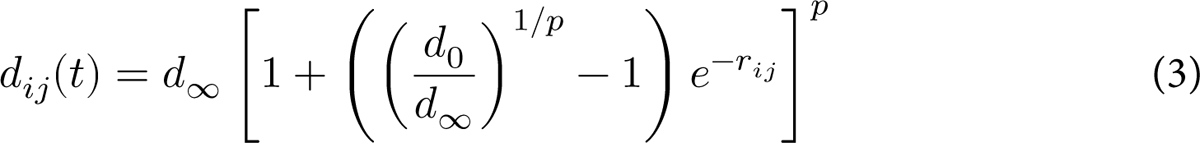

Given this equation, eq. (2) can be rearranged to allow the calculation of the lesion-specific growth rate at the growth inflection point. Eq. (4) is used to obtain by r_ij_ by substituting *d*_ij_(u) with 10, u_10ij_ with the lesion-specific value sampled from the Fréchet distribution.

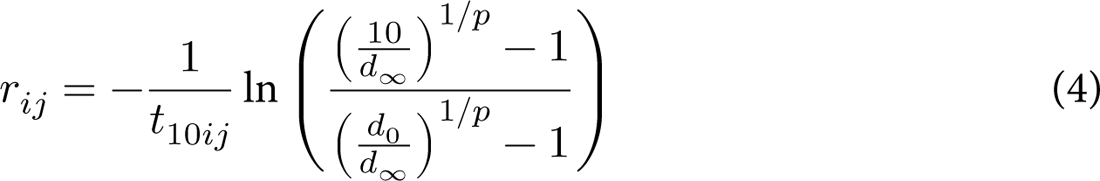

### 7.2 Transition from Adenoma to Cancer

The size at transition to preclinical cancer is modeled with a log-normal distribution, where the size of the jth adenoma of the ith person is defined as *ln(S_ij_)* ∼ *N*(μ_ij_, *σ*_γ_). The expected log-diameter at transition to preclinical cancer is defined by eq. (5).

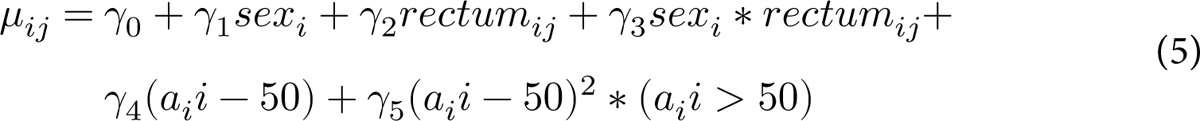

### 7.3 Time from Preclinical to Clinical Cancer (Soujourn Time)

Eq. (6) defines the soujourn time *T_ij_* (time from preclinical transition to symptomatic cancer) of adenomas. CRC-SPIN uses a Weibull distribution with scale parameter λ_1_, shape parameter λ_2_ and the λ_3_ parameter allows for different soujourn time for cancers in the colon and in the rectum.

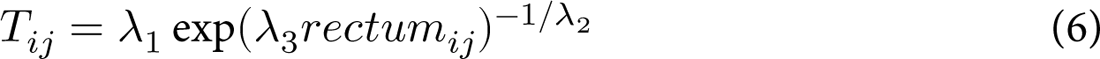

## 8 Appendix II: Colonoscopy Sensitivity

Colonoscopy sensitivity to detect adenomas conditional on their size is crucial in model-based screening cost-effectiveness analyses. The assumptions made about sensitivity directly define the expected effectiveness of screening strategies. Colonoscopy sensitivity is informed by tandem colonoscopy studies wherein two colonoscopies are performed on the same patient. Researchers count the number of adenomas missed by the first colonoscopy n_1_ and found in the second exam n_2_. The adenoma miss rate (AMR) is computed by the number of adenomas found in the second exam divided by the total number of adenomas found.

Existing studies (Zauber et al. 2008; Knudsen et al. 2016, 2021b) conventionally use data from tandem colonoscopy reviews (i.e., Van Rijn et al. 2006) to inform colonoscopy sensitivity assumptions. Nevertheless, the analysis provided by the Van Rijn et al. (2006) review might not be directly used to determine the expected colonoscopy sensitivity without further consideration. Miss rates reported in tandem colonoscopy studies or other designs offer an upper bound for colonoscopy sensitivity but not a lower bound.

This section develops a Bayesian approach to evaluate the plausibility of colonoscopy sensitivity scenarios conditional on adenoma size using the same data reviewed by Van Rijn et al. (2006) and assuming *sensitivity = 1 − AMR* results in a demonstrably biased estimator. This estimate can only reflect an upper bound for sensitivity under the assumption that the second exam missed no adenomas. Because about 1/4 of adenomas are missed in the first exam, this assumption is problematic. As diagnostic tests are improved, the “real” miss rate of conventional colonoscopy will gradually decline, predictably demonstrating that previously-made colonoscopy sensitivity assumptions were overly optimistic.

### 8.1 Data

Table 7 presents the list of studies used by Van Rijn et al. (2006). Each row represents one combination of adenoma size category and study included in the review. n1 is the number of adenomas found in the first colonoscopy, and n2 is the number found in the second exam. Adenoma miss rates *AMR* = n_2_/(n_1_ + n_2_) reflect the proportion of adenomas missed by the first exam and found in the second exam.

### 8.2 Model

Let *N* be the total number of adenomas present in a study population. The examiner counts the number of adenomas found in the first n_1_ and in the second exam n_2_, within a particular size category t ∈ [*d*, m, l], where *d* are diminutive adenomas (<5mm), m are medium adenomas (5-10mm) and l are large adenomas (>10mm). *N* might be close or equal to n_1_ + n_2_ if sensitivity is high, but *N* is never observed.

Eq. (7) describes a model of the data generation process underlying tandem colonoscopy studies, assuming the same, uncorrelated sensitivity for both exams. This model is conditioned on the size of the adenoma and represents a single study. Adenoma size subscripts are omitted from these equations for clarity.

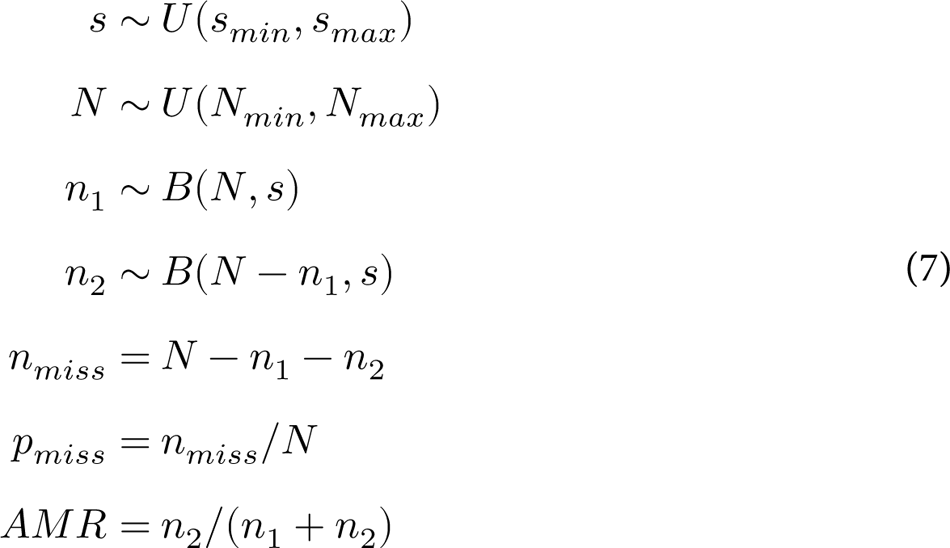

I specify uninformative priors for colonoscopy sensitivity S ranging from 0.05 to 1 and priors for *N* such that *N*_min_ = n_1_ + n_2_ and *N*_max_ = *N*_min_/(1 − p^w^_miss_)where *p^w^_miss_* 0.5 represents the maximum proportion of adenomas missed by both exams. Setting this proportion to 0 results in an illogical assumption if sensitivity is not 1, but is nevertheless an assumption made in prior studies. Setting it well above 0.5 would only be consistent with a pessimistic assumption that sensitivity is very low and adenoma prevalence is very high. Future analyses can plausibly use a more informative prior for *N* based on expected adenoma prevalence. However, we refrain from doing so in this analysis since the risk profile of the studies is heterogeneous, and known adenoma prevalence depends on the sensitivity of colonoscopy. Moreover, this appendix aims to provide plausible lower and upper bounds for sensitivity scenarios that are still consistent with data used by existing studies.

### 8.3 Plausibility of Sensitivity Assumptions

As stated, a known relationship exists between the unknown expected proportion of adenomas missed by two back-to-back colonoscopies *E*(p_miss_) and adenoma sensitivity S. We know that *E(p_miss_) = (1 − S)^2^ = 1 − 2S + S^2^*. If *S = 1*, then *E(p_miss_) = 0*. If *S = 0*, then *E(p_miss_) = 1*. If *S = 0.75*, then tandem colonoscopy studies will miss *E(p_miss_) = 1 − 2 ∗ 0.75 + 0.75^2^ = 0.065* of adenomas in expectation. If sensitivity is 50%, then these studies are expected to miss about 25% of adenomas in expectation. While tandem colonoscopy studies offer some information about sensitivity, this information is not sufficient to identify sensitivity.

To define bounds for sensitivity, we use the model outlined in eq. (7) to simulate each study reviewed by Van Rijn et al. (2006). The purpose is to derive plausible sensitivity values for the scenarios and illustrate what those sensitivity assumptions would imply about the proportion of adenomas missed by both exams. Figure 5 shows the posterior distribution for colonoscopy sensitivity to diminutive adenomas for a set of tandem studies and overlays the assumption made by each of the four sensitivity scenarios considered in this analysis as dotted lines. This figure demonstrates that the very high and baseline sensitivity scenario assumptions might be compatible with the sensitivity of Hixon, 1991. However, they are unlikely to have generated the results observed in Rex, 1997. The Very Low sensitivity assumption seems aligned with the mode of the posterior distribution of Harrison, 2004, and is not ruled out by results from Rex, 1997 and Rex, 2003 I. These results demonstrate that all sensitivity scenarios included in this study are plausible with at least one tandem colonoscopy study reviewed by Van Rijn et al. (2006) and should not be deemed implausible. Future work might extend this analysis to generate a full sensitivity distribution representing today’s colonoscopy studies.

**Figure 5:**
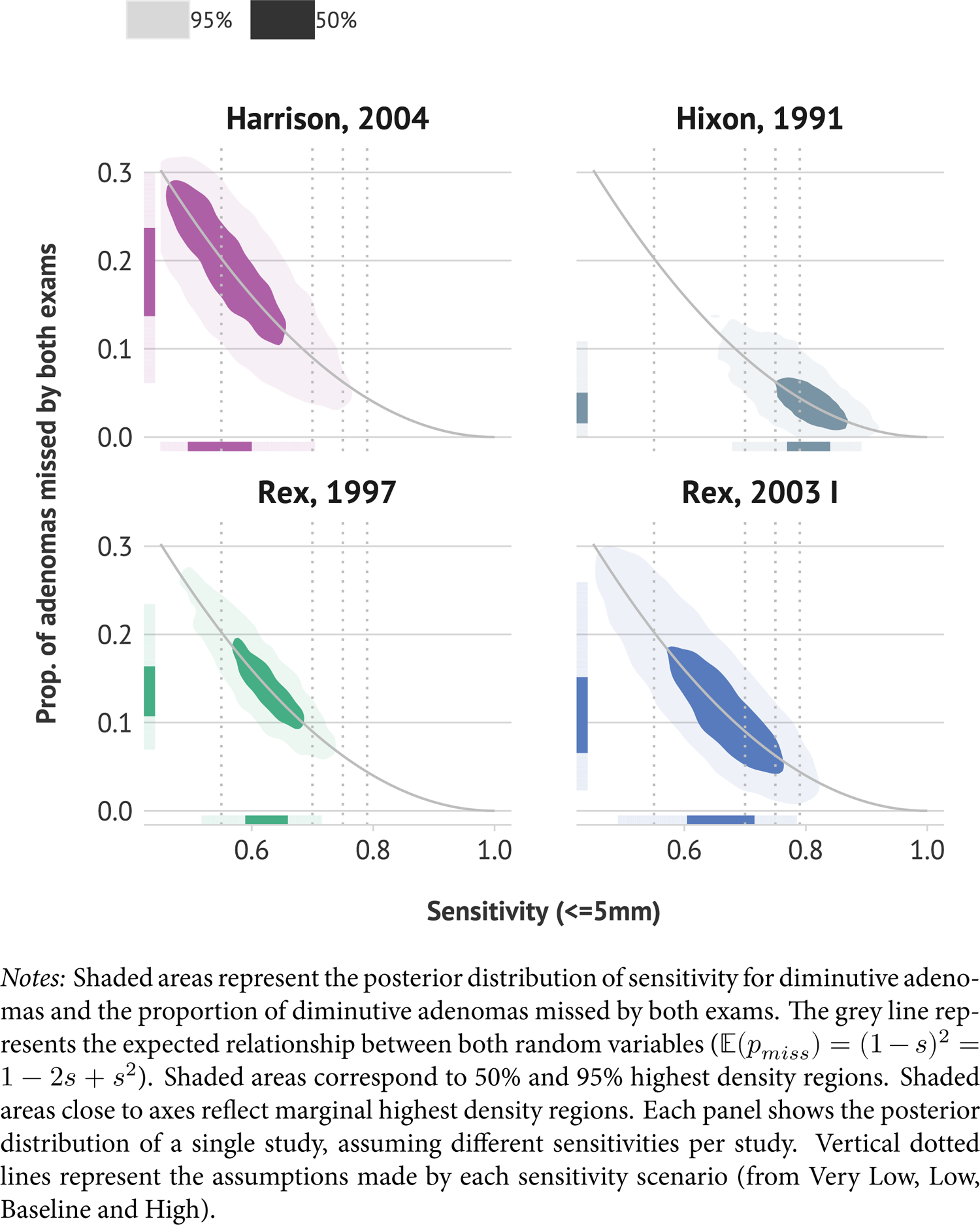
Posterior Distributions of Colonoscopy Sensitivity for Diminutive Adenomas (less than 5 mm) for Select Tandem Colonoscopy Studies

### 8.4 Continuous-size Sensitivity Functions

While table 3 presents average sensitivity conditional on categorical sizes, CRC-SPIN is a continuous-size, continuous-time model and requires sensitivity to be defined for any arbitrary adenoma size between 1 and 50 mm. In prior analyses, CRC-SPIN has used a quadratic functional form to approximate the baseline sensitivity. Instead, this analysis uses the Stineman interpolation method Jhannesson, Bjornsson, and Grothendieck (2018) to generate plausible sensitivity assumptions consistent with the categorical average sensitivities. The Stineman interpolation method offers a few advantages over the prior quadratic functional form, allowing direct control over the minimum and maximum sensitivity values, more flexibility over the functional form of the sensitivity curve, and ensuring that sensitivity is monotonic. Figure 6 shows the resulting sensitivity functions produced with the monotonic Stineman interpolation method. These sensitivity curves match the average categorical sensitivities in table 3.

**Figure 6:**
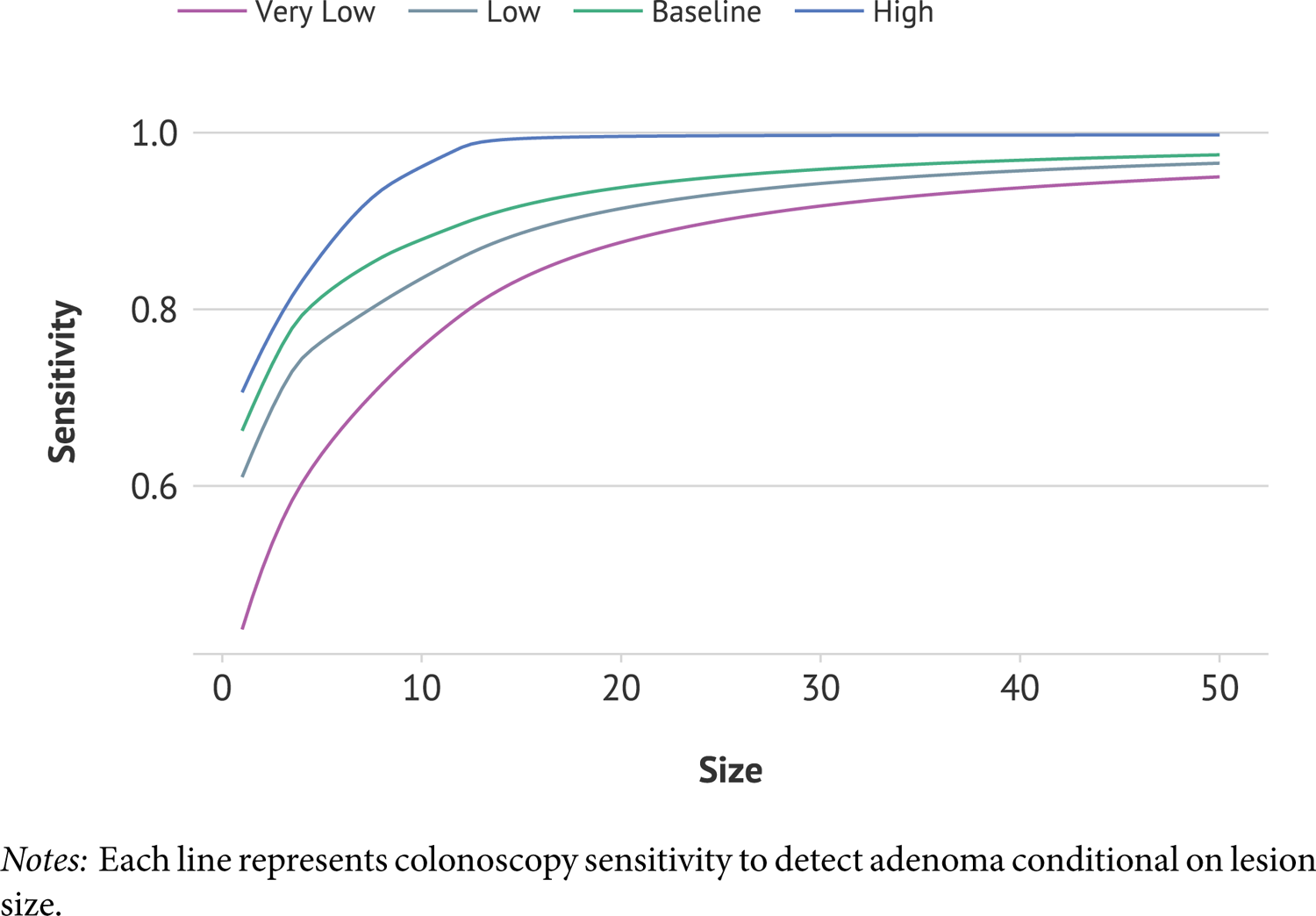
Continuous Colonoscopy Sensitivity Functions by Scenario

## 9 Appendix III: Computing Environment

Model calibration and screening strategy experiments were performed on Bebop and Theta at Argonne National Laboratory. Bebop is a High-Performance Computing cluster managed by the Laboratory Computing Resource Center at Argonne National Laboratory. This study used approximately 200,000 core hours to calibrate both model specifications and around 200,000 core hours to run screening experiments. Four HPC workflows were developed to support this study. All workflows were based on Swift-T (Wozniak et al. 2013) and EMEWS (Ozik et al. 2016).

## 10 Appendix IV: The crcrdm R package

The crcrdm R package (Nascimento de Lima 2022) was designed to support this and future large-scale microsimulation studies. crcrdm is an R package created to support large-scale cancer screening analyses based on cancer microsimulation models. The package supports large-scale computational tasks historically deemed unfeasible for microsimulation models, such as defining and conducting Probabilistic Sensitivity Analyses (PSAs) or robustness analyses of large models and large combinations of parameter sets. The package helps the user perform large-scale experiments with microsimulation models by partitioning the memory usage of models to a manageable size and organizing the experimental design to run across different nodes (e.g., the same model run is parallelized across different computers). The package also supports multi-model experimental designs. This package implements the crcmodel and the crcexperiment classes and can be used to perform Robust Decision Making Analyses of multiple cancer screening models using high-performance computing resources.

## Footnotes

1 The Stineman method (Stineman 1980; Jhannesson, Bjornsson, and Grothendieck 2018) provides a well-behaved smooth interpolation approach. We defined a set of year knots at which the birth cohort effect is estimated and used the Stineman interpolation method for the years between the knots. Unlike methods based on polynomials such as splines, the Stineman interpolation is guaranteed to be monotonic.

2 Following Knudsen, et al. (2021a), we do not use the term Incremental Cost-Effectiveness ratios because this analysis does not consider costs, but the number of colonoscopies as a proxy for the burden of screening.

3 We verified that doubling the sample size does not change any of the estimates presented in this paper.

4 This figure presents a low-dimensional version of what would otherwise be one of the outputs of RDM’s vulnerability analysis step. Note that colonoscopy sensitivity could be expressed by a continuous variable. We choose colonoscopy sensitivity scenarios that match the values used in existing analyses to allow comparisons to the existing literature. We add a new “Very Low” sensitivity scenario that we show is plausible in Appendix

